# Phenome-wide association study of *TTR* and *RBP4* genes in 361,194 individuals reveals novel insights in the genetics of hereditary and senile systemic amyloidoses

**DOI:** 10.1101/19001537

**Authors:** Antonella De Lillo, Flavio De Angelis, Marco Di Girolamo, Marco Luigetti, Sabrina Frusconi, Dario Manfellotto, Maria Fuciarelli, Renato Polimanti

**Author notes:** Corresponding author: Dr. Renato Polimanti, Department of Psychiatry, Yale University School of Medicine and VA CT Healthcare, Center, VA CT 116A2, 950 Campbell Avenue, West Haven, CT 06516, USA.

## Abstract

*Transthyretin* (*TTR*) gene has a causal role in a hereditary form of amyloidosis (ATTRm) and is potentially involved in the risk of senile systemic amyloidosis (SSA). To understand the genetics of ATTRm and SSA, we conducted a phenome-wide association study of *TTR* gene in 361,194 participants of European descent testing coding and non-coding variants. Among the 382 clinically-relevant phenotypes tested, *TTR* non-coding variants were associated with 26 phenotypic traits after multiple testing correction. These included signs related to both ATTRm and SSA such as chronic ischaemic heart disease (rs140226130, p=2.00×10^−6^), heart failure (rs73956431, p=2.74×10^−6^), atrial fibrillation (rs10163755, p=4.63×10^−6^), dysphagia (rs2949506, p=3.95×10^−6^), intestine diseases (rs970866, p=7.14×10^−6^) and anxiety (rs554521234, p=8.85×10^−6^). Consistent results were observed for *TTR* disease-causing mutation Val122Ile (rs76992529) with respect to carpal tunnel syndrome (p=6.41×10^−6^) and mononeuropathies of upper limbs (p=1.22×10^−5^). Sex differences were also observed in line with ATTRm and SSA epidemiology. Additionally, we explored possible modifier genes related to TTR function, observing convergent associations of *RBP4* variants with the clinical phenotypes associated with *TTR* locus. In conclusion, we provide novel insights regarding the molecular basis of ATTRm and SSA based on large-scale cohort, expanding our understanding of the phenotypic spectrum associated with *TTR* gene variation.

## INTRODUCTION

Transthyretin (TTR) misfolding and the consequent amyloid formation and deposition lead to two form of amyloidoses: a hereditary (or mutant) form (ATTRm) caused by more than 100 *TTR* gene coding mutations [1], and a senile systemic form (SSA) that is due to the misfolding of wild type protein (i.e., no *TTR* coding mutation present in the individuals [2]. ATTRm is a life-threating disorder that affects several tissues (e.g. brain, autonomic and peripheral nerves, heart, and gastrointestinal (GI) tract) and is characterized by an extreme heterogeneity in the genotype-phenotype correlation, including the age of onset, the penetrance, and the clinical display [3-6]. Despite *TTR* coding mutations are the cause of the disease, the clinical variability of the ATTRm is likely due to several other factors [7-11]. Differently from ATTRm, patients affected by SSA do not present a coding mutation in *TTR* gene and the main symptoms include cardiac failure and carpal tunnel syndrome[12-16]. *TTR* non-coding variation via its regulatory function seems to have a role in the phenotypic heterogeneity of ATTRm and to play a role in the pathogenesis of SSA [11,17-19]. Furthermore, variation in other genes encoding for protein products that interact with TTR tetramer seems to modulate the pathogenetic processes involved in the TTR fibrils formation and, consequently, in the course of the disease [20-22]. Genome-wide datasets generated from cohorts including hundred thousand participants can be leveraged to disentangle complex genotype-phenotype associations. In particular, phenome-wide association studies (PheWAS) can permit us to detect novel associations with respect to known risk loci with respect to a wide range of phenotypes [23-25]. In the present study, we explored the phenotypic spectrum associated with the coding and non-coding variants of *TTR* gene, also investigating potential modifier loci that could modulate TTR amyloidogenic process. To date, the vast of majority of the studies that investigated the genetics of ATTRm and SSA have been conducted on cohorts with a limited sample size due to the low disease prevalence [9,11,19]. ATTRm and SSA signs are poorly recognized and the correct diagnosis in sporadic cases is usually established several years from the onset of the symptoms [26]. This consistently limited the ability to collect large informative cohorts needed to investigate the genetics of ATTRm and SSA. To date, it is possible to investigate cohorts including hundred thousand participants to explore the association of genetic variation with clinically relevant phenotypes. Accordingly, we conducted a PheWAS in 361,194 participants of European descent available from the UK Biobank with respect to 382 clinically relevant traits to investigate genetic variation potentially involved in TTR-related pathogenic processes. The results obtained support that non-coding variants located in *TTR* gene and other disease-modifying loci are associated with phenotypic traits related to ATTRm and SSA. On the basis of the basis of the current findings, we hypothesize that non-coding variation is involved i) in the variability of genotype-phenotype correlation of carriers of *TTR* coding mutations; and ii) in the increased risk of SSA in non-carriers of *TTR* coding mutations.

## MATERIALS AND METHODS

### Genome-wide datasets

The dataset used for the analysis was derived from the UK Biobank. This is an open access resource available to investigate a wide range of serious and life-threatening illnesses [27]. This project has recruited more than 500,000 people assessed for a wide range of phenotypic information, also including clinically-relevant phenotypes. Complete genome-wide data are available for the whole cohort. These genetic data were used to generate genome-wide association datasets that can be used to explore the genetics of human diseases and traits. These genome-wide datasets used in the present study were generated on the analysis of 361,194 participants of European descent including 194,174 women and 167,020 men. The association analysis for all phenotypes was conducted using a linear regression model available in Hail (available at https://github.com/hail-is/hail) including the first 20 ancestry principal components, sex, age, age^2^, sex×age, and sex×age^2^ as covariates. Details regarding QC criteria, GWAS methods, and the original data are available at https://github.com/Nealelab/UK_Biobank_GWAS/tree/master/imputed-v2-gwas.

### Clinically-relevant Phenotypes

Since our goal was to investigate phenotypes related to the pathogenesis of ATTRm and SSA, we focused on clinically-relevant traits available in the UK Biobank. Specifically, we considered ICD-10 (International Classification of Disease, 10th Revision; [28,29]) codes and clinical endpoints derived by the FinnGen project [https://www.finngen.fi/en]. ICD-10 codes are a specific terminology that include diseases, signs, symptoms, and procedure codes maintained by the World Health Organization (WHO), and they typically are used to billing data [28]. The FinnGen project comes from a collaboration among Finnish universities, biobanks, hospital districts, and several international pharmaceutical companies to increase knowledge about the origins of diseases. FinnGen clinical endpoints (available at https://www.finngen.fi/en/researchers/clinical-endpoints) were developed to conduct genome-wide investigations of phenotypic traits assessed across several national health registers. To remove phenotypes not informative due to a lack of statistical power, we investigated phenotypic traits with a number of cases greater than 1000 individuals. The full list of the phenotypes investigated and their corresponding sample size is reported in the Table S1. Additionally, we also conducted sex-stratified analysis to investigate the known differences between sexes in the epidemiology of ATTRm and SSA. A total of 382, 240, and 225 clinically-relevant phenotypes were investigated in the total sample and in the sex-stratified analysis (female- and male-specific analyses, respectively).

### Data Analysis

We considered a total of 382 clinically-relevant phenotypes (sex-stratified analysis: 240 for female-specific PheWAS and 225 for male-specific PheWAS) and investigated variability of *TTR* gene considering a 4-Mb region (NC_000018.9: 27,171,000–31,171,500) including the genic locus (NM_000371; 7,257 bp) and the surrounding regions (± ∼2Mb from the transcription start/end sites) (Table 1). We only considered variants with a minor allele frequency (MAF) greater than 1%. As recommended in the original UK Biobank analysis (information available at http://www.nealelab.is/blog/2017/9/11/details-and-considerations-of-the-uk-biobank-gwas), we considered high-confidence association results generated from variants with at least 25 minor alleles in the smaller group (case or control). A total of 12,719, 12,710, and 12,711 high-confidence *TTR* variants were investigated in the overall-sample and in the sex-stratified analyses (male- and female-specific, respectively). False Discovery Rate (FDR) at 10% was applied as significance threshold for multiple testing correction [30] accounting for the number of variants and the number of traits tested. To estimate the independent association signals within the genomic region tested, PLINK 1.09 [31] was used to perform linkage disequilibrium (LD) clumping considering a 0.1 R^2^ cut-off within a 500-kb window.

**Table 1:** Details regarding the clinically-relevant traits and the genomic regions tested in the overall cohort and in the sex-stratified analyses.

| Gene | Localization | Sex | Phenotype N | Case Median N (Min – Max) | Control Median N (Min-Max) |
| --- | --- | --- | --- | --- | --- |
| *TTR* | NC_000018.9:<br>27,171,000 – 31,171,500 | BOTH SEXES | 382 | 2,534 (1011 – 208,009) | 358,660 (79,185 – 360,183) |
|  |  | FEMALE | 240 | 2,364 (1,008 – 153,946) | 191,810 (40,228 – 193,166) |
|  |  | MALE | 225 | 2,533 (1,004 – 128,063) | 164,487 (39,957 – 166,016) |
| *RBP4* | NC_000010.10:<br>93,353,000 – 97,353,500 | BOTH SEXES | 32<sup>a</sup> | 2,820 (1,014 – 20,857) | 358,374 (340,337 – 360,180) |
|  |  | FEMALE | 32<sup>a</sup> | 1,095 (331 – 7,355) | 193,079 (186,819 – 193,843) |
|  |  | MALE | 31<sup>a</sup> | 1,581 (441 – 15,056) | 165,439 (151,964 – 166,579) |

<sup>a</sup>significant phenotypic traits identified in the *TTR* PheWAS and tested in the *RBP4* analysis.

STRING v.11.0 [32] was used to identify protein interaction with *TTR*, considering experiments, co-expression, co-occurrence, gene fusion, and neighbourhood as active sources and a confidence score higher than 0.9. With respect to proteins identified, we investigated variants included in coding, non-coding, and surrounding regions of the encoding gene using the same quality control criteria applied in the analysis of *TTR* variants. Additionally, we investigated the functional enrichments association related to the protein-protein interactions identified considering Gene Ontologies [33,34] for biological processes and molecular functions and molecular pathways available in the Reactome Database [35].

## RESULTS

The *TTR* PheWAS identified several phenotypic associations surviving FDR multiple testing correction (Figure 1). These include associations identified in the overall analysis and in the sex-stratified analyses (Figure 2, Table S2). In the analysis of the total cohort (Table S3), we observed several clinical signs that may be related to ATTRm and SSA: spinal stenosis (FinnGen: M13_SPINSTENOSIS; rs116937108, beta=0.002, p=1.21×10^−7^), neuromuscular dysfunction of bladder (ICD-10: N31; rs9953311, beta=-0.001, p=2.30×10^−6^; rs575854233, beta=-0.001, p=5.33×10^−6^), and anxiety disorders (FINNGEN: KRA_PSY_ANXIETY; rs554521234, beta = 0.003, p=8.85×10^−6^). Additionally, we also observed significant findings not expected to be directly related to TTR amyloidogenic process: Barret’s esophagus (FinnGen: K11_BARRET; rs147570462, beta=0.003, p=1.19×10^−7^), abscess of anal and rectal regions (ICD-10: K61; rs72948149, beta=0.002, p=1.87×10^−6^); fissure and fistula of anal and rectal regions (ICD-10: K60; rs72927361, beta=-0.003, p=1.90×10^−6^); complications of other internal prosthetic devices, implants and grafts (ICD-10: T85; rs34444556, beta=0.003, p=5.5×10^−6^), ulcer of esophagus (FinnGen: K11_OESULC; rs74481699, beta=0.004,p=6.43×10^−6^), adjustment and management of implanted device (ICD-10: Z45; rs117672477, beta=0.002, p=6.51×10^−6^); primary lymphoid and hematopoietic malignant neoplasms (FinnGen: C3_PRIMARY_LYMPHOID_HEMATOPOIETIC; rs17718949, beta=0.01, p=6.57×10^−6^). Among the 240 clinically-relevant phenotypes tested in the female-specific analysis (N = 194,174), we identified 8 LD-independent variants associated with specific pathological conditions (Table S4): chronic ischaemic heart disease (ICD-10: I25; rs140226130, beta=0.003, p=2.00×10^−6^), dysphagia (ICD-10: R13; rs2949506, beta=0.002, p=3.95×10^−6^), other disorders of urinary system (ICD-10: N39; rs71173870, beta=0.003, p=9.81×10^−6^), other specified/unspecified soft tissue disorders (ICD-10: M13_SOFTTISSUENAS; rs558461933, beta=0.007, p=9.92×10^−6^), other cataract (ICD-10: H26; rs12966815, beta=0.003, p=1.10×10^−5^), retinal detachments and breaks (ICD-10: H33; rs117637258, beta=0.003, p = 1.12×10^−5^), other abnormal products of conception (ICD-10: O02; rs139327590, beta=0.004, p=1.16×10^−5^), and shoulder lesions (ICD-10: M75; rs77728273, beta=-0.004, p=1.21×10^−5^). Several of these clinical manifestations (e.g., I25∼Chronic ischaemic heart disease, R13∼Dysphagia, N39∼Other disorders of urinary system, H26∼Other cataract and H33∼Retinal detachments and breaks) seem to be closely related to the expected symptoms of ATTRm and SSA. The male-specific PheWAS (225 phenotypic traits tested) revealed several significant associations that could be related to cardiac, gastrointestinal, and urinary symptoms of ATTRm and SSA (Table S5): other disorders of bladder (ICD-10: N32; rs138038371, beta=0.009, p=5.01×10^−7^), heart failure (FinnGen: I9_HEARTFAIL; rs73956431, beta=0.005, p=2.74×10^−6^), malignant neoplasm of colon (ICD-10: C18; rs78431500, beta=0.006, p=2.89×10^−6^), Barret’s esophagus (ICD-10: K11_BARRET; rs147570462, beta=0.005, p=3.45×10^−6^), atrial fibrillation and flutter (ICD-10: I48; rs10163755, beta=0.003, p=4.63×10^−6^), calculus of kidney and ureter (ICD-10: N20; rs8090264, beta=0.002, p=5.23×10^−6^), complications of procedures not elsewhere classified (ICD-10: T81; rs182526571, beta=0.008, p=6.57×10^−6^), other diseases of intestine (ICD-10: K63; rs970866, beta=-0.005, p=7.14×10^−6^) and fibroblastic disorders (FinnGen: M13_FIBROBLASTIC; rs60487427, beta=-0.004, p=8.38×10^−6^).

**Figure 1:**
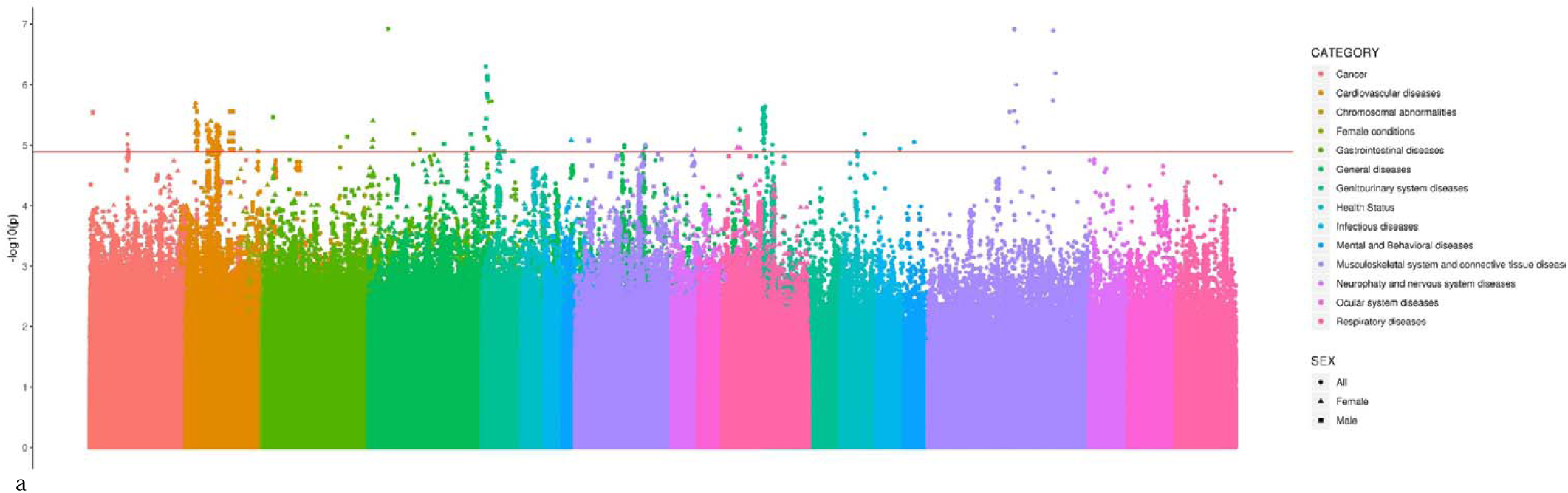
Manhattan Plot of *TTR* PheWAS in the total sample and in sex-stratified analyses across the different phenotypic categories tested.

**Figure 2:**
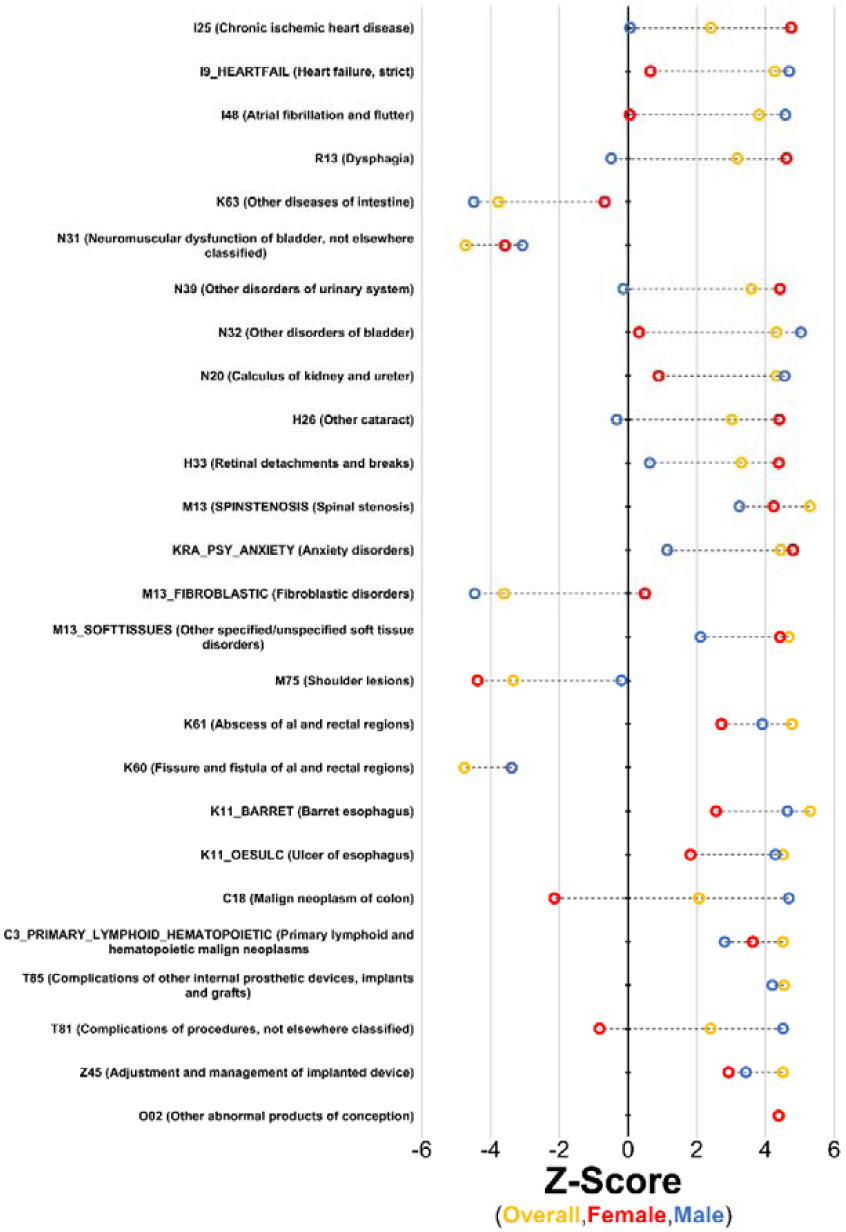
Significant associations of *TTR* non-coding variants with clinically-relevant phenotypes in the total sample and in the sex-stratified analyses. The full details of the associations are reported in Table S2.

Amyloidogenic coding variants in *TTR* gene are the established cause of ATTRm [1,5,6]. The large sample size of the UK Biobank cohort includes carriers of *TTR* amyloidogenic mutations with high-quality genotype information (imputation info score > 0.8; Table S6). However, due to the very low MAF of these coding mutations, the phenotypic associations of these variants should be considered only a “low-confidence” result as described in the methods. Being aware of this limitation, we explored their phenotypic spectrum in the UK Biobank (Table 2). The most relevant results were observed for the *TTR* Val122Ile mutation (rs76992529) in the female participants with respect to two well-known symptoms of the amyloidogenic process: carpal tunnel syndrome (FinnGen: G6_CARPTU; beta=0.307, p=6.41×10^−6^) and mononeuropathies of upper limb (ICD-10: G56; beta=0.306, p=1.22×10^−5^).

**Table 2:** Significant associations identified with respect to *TTR* coding mutations. Information about beta value, se, p-value and FDR q-value are reported. Bold text: TTR protein substitution. Underline text: TTR precursor substitution. Information about allele frequency and minor allele frequency are reported in Table S9.

| PHENOTYPIC TRAIT |  | RSID | SUBSTITUTION | BETA | SE | P-VALUE | FDR |
| --- | --- | --- | --- | --- | --- | --- | --- |
| Both sexes | FinnGen: XVII_MALFORMAT_ABNORMAL~Congenital malformations, deformations and chromosomal abnormalities | rs138657343 | <u>Arg5His</u> | 0.0264 | 0.00550 | $1.64 \times 10^{-6}$ | 0.029 |
| Female | ICD10: R87~Abnormal findings in specimens from female genital organs | rs138657343 | <u>Arg5His</u> | 0.0379 | 0.00724 | $1.65 \times 10^{-7}$ | 0.004 |
| | FinnGen: XVII_MALFORMAT_ABNORMAL~Congenital malformations, deformations and chromosomal abnormalities | | | 0.0368 | 0.00756 | $1.12 \times 10^{-6}$ | 0.019 |
| | ICD10: N94~Pain and other conditions associated with female genital organs and menstrual cycle | rs76992529 | **Val122Ile** | 0.158 | 0.0334 | $2.31 \times 10^{-6}$ | 0.032 |
| | FinnGen: G6_CARPTU~Carpal tunnel syndrome | | | 0.307 | 0.0681 | $6.41 \times 10^{-6}$ | 0.063 |
| | ICD10: G56~Mononeuropathies of upper limb | | | 0.306 | 0.0699 | $1.22 \times 10^{-5}$ | 0.079 |
| | FinnGen: K11_HERNIA~Hernia | rs121918074 | **His90Asn** | 0.210 | 0.0485 | $1.53 \times 10^{-5}$ | 0.090 |

To test the presence of modifier genes involved in TTR-related pathogenic processes, we investigated potential protein interaction and observed the highest-confidence interaction with respect to RBP4 protein, which is is supported by experiments and co-expression interaction sources (STRING interaction score = 0.914). The TTR-RBP4 protein interaction is associated with functional enrichments for GO terms, including retinol metabolic process (GO:0042572, FDR q=6.3×10^−4^) and protein heterodimerization activity (GO:0046982, FDR q=0.02), and for Reactome molecular pathways, including retinol cycle disease events (HSA-2453864, FDR q=9.98×10^−6^), canonical retinoid cycle in rods (HSA-2453902, FDR q=1.1×10^−5^), and retinoid metabolism and transport (HSA-975634, FDR q=2.84×10^−5^). On the basis of these data, we hypothesize a convergence where variants located in *TTR* and *RBP4* genes are associated with clinically-relevant phenotypes related to ATTRm and SSA pathogenesis. Accordingly, we investigated 13,226, 13,231, and 13217 high-confidence variants (overall-sample, female-specific, and male-specific analyses, respectively) located in *RBP4* gene and its surrounding regions (NC_000010.10: 93,353,000 – 97,353,500) with respect to the significant phenotypic traits observed in the *TTR* PheWAS (Table 1). We identified significant associations in the overall cohort and in the sex-stratified analyses (Table S2). In the overall sample, we confirmed some of the *TTR* associations previously observed with respect to *TTR* gene that are related to known clinical signs of ATTRm and SSA (Table S7): heart failure (FinnGen: I9_HEARTFAIL; rs1326222, beta=0.001, p=5.25×10^−7^) and anxiety disorders (FinnGen: KRA_PSY_ANXIETY; rs112059561, beta=0.002, p=6.44×10^−6^). Among the phenotypes that do not seem to be related to TTR amyloidogenic processes, Barret’s esophagus was also significantly associated with a RBP4 variant (FinnGen: K11_BARRET; rs12573026, beta=0.001, p=9.96×10^−8^). In the female-specific analysis, we observed significant associations with respect to cardiac symptoms which are one of the leading sigsn of ATTRm and SSA: atrial fibrillation and flutter (ICD-10: I48; rs4917692, beta=0.002, p=1.15×10^−5^) and heart failure (FinnGen: I9_HEARTFAIL; rs1326222, beta=0.001, p=1.23×10^−5^) (Table S8). Finally, in the male participants, we observed a convergence between *RBP4* and *TTR* findings with respect to two additional phenotypes expected to be related to ATTRm and SSA (FinnGen: M13_SPINSTENOSIS∼spinal stenosis, rs112288944, beta=0.005, p=1.42×10^−5^; ICD-10: R13∼Dysphagia, rs61886346, beta=0.003, p=1.66×10^−5^). Some conditions that are not expected to be linked to amyloidogenic processes such as Barret’s esophagus (FinnGen: K11_BARRET; rs12573026, beta=0.002, p=1.01×10^−7^) and malignant neoplasm of colon (ICD-10: C18; rs142083973, beta=0.005, p=3.40×10^−6^) were also associated with both *RBP4* and *TTR* gene variants in the male sample (Table S9). Table S10 summarizes all significant associations, providing the allele frequencies of *TTR* and *RBP4* variants identified.

## DISCUSSION

TTR misfolding and the following amyloid formation and deposition are the cause of ATTRm and SSA due to *TTR* coding variants and the misfolding of the wild type protein, respectively [1,2]. With respect to ATTRm, several studies investigated the role of *TTR* coding and non-coding variation to dissect the molecular machineries at the basis of its complex phenotype-genotype correlation [8-11, 17,18, 19]. Differently, limited information is available regarding the genetic basis of SSA [19]. PheWAS design is a powerful tool to broaden the knowledge about the phenotypic spectrum associated with disease-causing genetic variations [23-25] and its performance is enhanced by the availability of genomic data of large cohorts. Accordingly, we conducted a phenome-wide investigation of *TTR* coding and non-coding variants in more than 300,000 participants of European descent, also evaluating the *RBP4* gene as a possible modifier of *TTR* pathogenetic mechanisms.

Our results pointed out novel associations of *TTR* non-coding variants with phenotypic conditions potentially related to ATTRm and SSA pathogenesis. In the sex stratified analysis, we observed strong evidence of the effect of *TTR* non-coding variations on cardiac involvement (ICD-10: I25∼Chronic ischaemic heart disease; I48∼Atrial fibrillation and flutter; FinnGen: I9_HEARTFAIL∼Heart failure, strict), one of the leading clinical signs of ATTRm and SSA. With respect to SSA, heart is the most affected organ in elderly patients, and, the main manifestations are cardiomyopathy (resulting in increase of biventricular wall thickness and ventricular stiffness) and atrial fibrillation [26,36]. The symptoms associated with ATTRm include a restrictive amyloid cardiomyopathy along with a combination of several other signs, including gait, gastrointestinal, neurological, urinary/renal, and ocular involvement frequently reported among the affected carriers [1,6,26,36].

GI symptoms were identified in our phenome-wide investigation (ICD-10: R13∼Dysphagia and K63∼Other diseases of intestine). The occurrence of GI manifestations has been reported in both ATTRm and SSA and these include nausea, vomiting, constipation, faecal incontinence, and weight loss [26,37,38]. Although GI involvement is reported in SSA patients, these symptoms are less common than the ones reported in ATTRm [38], where GI manifestations are more frequent in early-onset patients [38].

Significant associations of *TTR* variants with respect to bladder, urinary tract and kidney were identified in the overall sample and in the sex-stratified analyses (ICD-10: N31∼Neuromuscular dysfunction of bladder, not elsewhere classified; N39∼Other disorders of urinary system; N32∼Other disorders of bladder; N20∼Calculus of kidney and ureter). It is well known the evolution of neurogenic bladder in ATTRm that is linked to autonomic nerve dysfunction, and, renal involvement due to the amyloid deposition in the glomeruli arterioles and medium vessels [26,39].

The female sample showed that *TTR* non-coding variants are associated with ocular involvement (ICD-10: H26∼Other cataract; H33∼Retinal detachments and breaks). These findings are consistent with the known ocular manifestations described in ATTRm, which, in agreement with our sex-stratified result, occur more frequently in women [40]. The ocular symptoms reported in ATTRm patients include vitreous opacities, retinal vein occlusion and direct optic nerve infiltration[40].

Remarkably, a significant genetic association between *TTR* gene and anxiety disorders (FinnGen: KRA_PSY_ANXIETY) was observed. There is a growing literature regarding the effect of rare life-threatening diseases on the mental health of the patients and their families. With respect to ATTRm, several studies reported considerable psychological consequences in carriers of *TTR* mutations after the onset of the symptoms and the diagnosis of the disease [41,42]. These behavioural changes seem to be related to the stressful scenarios related to amyloidosis diagnostic path, including the pre-symptomatic genetic testing confirming the presence of an amyloidogenic mutation in family members and the time (usually years) intervening between the onset of the symptoms and the diagnosis of the diseases in novel ATTRm cases.

Spinal stenosis (FinnGen: M13_SPINSTENOSIS) was another phenotype identified in our *TTR* Phewas. This symptom has been reported in both ATTRm and SSA patients [26,36], and it usually co-occurs with other known signs of *TTR* amyloidogenic process such as carpal tunnel syndrome and neuropathies [43]. These phenotypic traits were identified as associated to *TTR* Val122Ile mutation (Table 2). This is also consistent with phenotypic presentation of Val122Ile carriers affected by ATTRm where the carpal tunnel syndrome represents an early sign of the disease whereas upper limb involvement is a clinical manifestation observed in a later stage [26]. Although our findings are related to carriers of European descent and Val122Ile mutation is mainly identified in individuals of African descent, a similar clinical phenotype has been reported in Val122Ile carriers of both ancestry groups [44].

Our findings support a phenotypic convergence between *TTR* and *RBP4* genetic associations, also including traits expected to be associated with ATTRm and SSA clinical spectrum (i.e., heart failure and atrial fibrillation). This evidence supports the putative role of *RBP4* as modifier gene with respect to TTR amyloidogenic process. RBP4 interacts with TTR via formation of TTR-RBP4 protein complex that stabilizes the TTR tetramer, inhibiting monomer dissociation and fibril formation [3]. In carriers of *TTR* amyloidogenic mutations, there is an increase of the TTR tetramer dissociation with a consequent decrease of the TTR-RBP4 complex and an increase of the RBP4 urinary excretion [3,45]. Accordingly, *RBP4* genetic variation could affect the functionality of the TTR-RBP4 complex, contributing to the complex phenotype-genotype correlation in carriers of *TTR* amyloidogenic mutations and increasing the risk of SSA in non-carriers.

Both *TTR* and *RBP4* association analyses identified traits that are not expected to be related to the pathological consequences of the amyloidogenic process. However, they may be related to the inter-individual variability of the main physiological function of these two genes: the transport of retinol (vitamin A) [46]. Altered homeostasis of vitamin is associated with several disorders [47] and some of them could be associated with the effect of *TTR* and *RBP4* variants on retinol metabolism, explaining some of the phenotypic traits identified in the present study.

In conclusion, the present study provides novel insights about *TTR* gene variation, confirming its role in phenotypic variability observed in ATTRm patients and supporting its potential involvement in the predisposition to SSA. These data support the relevance of large biobanks to investigate complex genotype-phenotype associations. Additionally, the insights provided support the necessity of further investigations of *TTR* non-coding variation together with putative modifier loci to develop tools able to anticipate the course of the disease in order to improve the diagnosis and management of patients affected by ATTRm and SSA.

## Data Availability

All data generated during this study are included in this published article and its Supplemental information files

## LIST OF ABBREVIATIONS

ATTRm: transthyretin-related hereditary (or mutant) form of amyloidosis
FDR: false discovery rate
GI: gastrointestinal
ICD-10: International Classification of Diseases 10th Revision
MAF: minor allele frequency
PheWAS: phenome wide association study
RBP4: retinol-binding protein 4
SSA: senile systemic amyloidosis
TTR: transthyretin
WHO: World Health Organization

## DECLARATIONS

### Ethics approval and consent to participate

Owing to the use of previously collected, de-identified, aggregated data, this study did not require institutional review board approval.

### Competing Interests

Drs. Fuciarelli and Polimanti are both receiving research grants from Pfizer Inc. to conduct epigenetic studies of TTR amyloidosis. The other authors reported no biomedical financial interests or potential conflicts of interest.

### Funding

This work was partially supported by the PhD Program of the University of Rome Tor Vergata (ADL) and the Yale University School of Medicine (RP).

### Author contributions

ADL and RP were involved in study design. ADL and RP carried out statistical analysis. All authors were involved in the interpretation of the results. ADL and RP wrote the first draft of the manuscript and all authors contributed to the final version of the manuscript.

## Acknowledgments

We thank the participants and investigators of the UK Biobank and the Neale lab for generating the genome-wide data used in the present study.

---

**Table S1:**
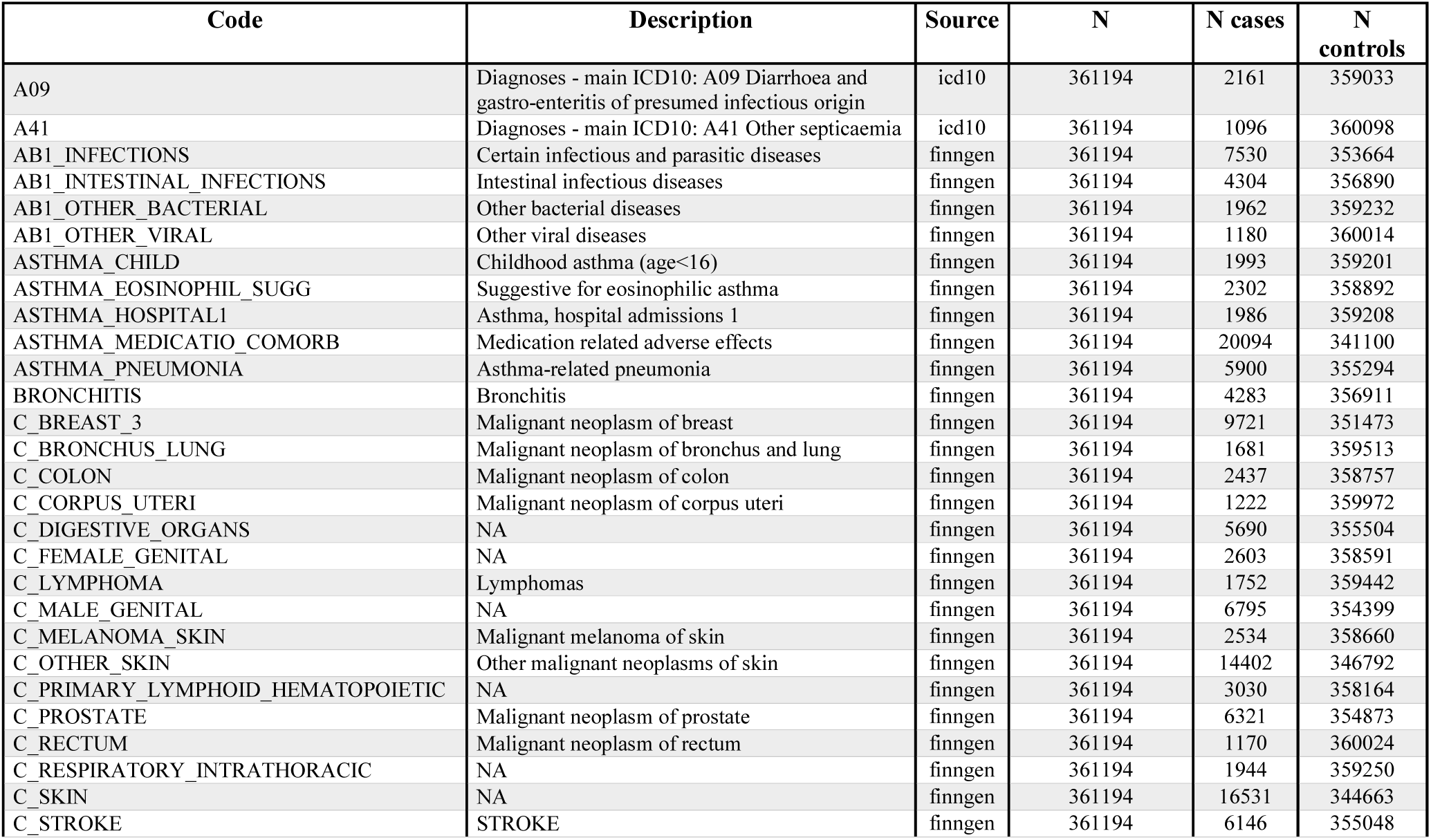
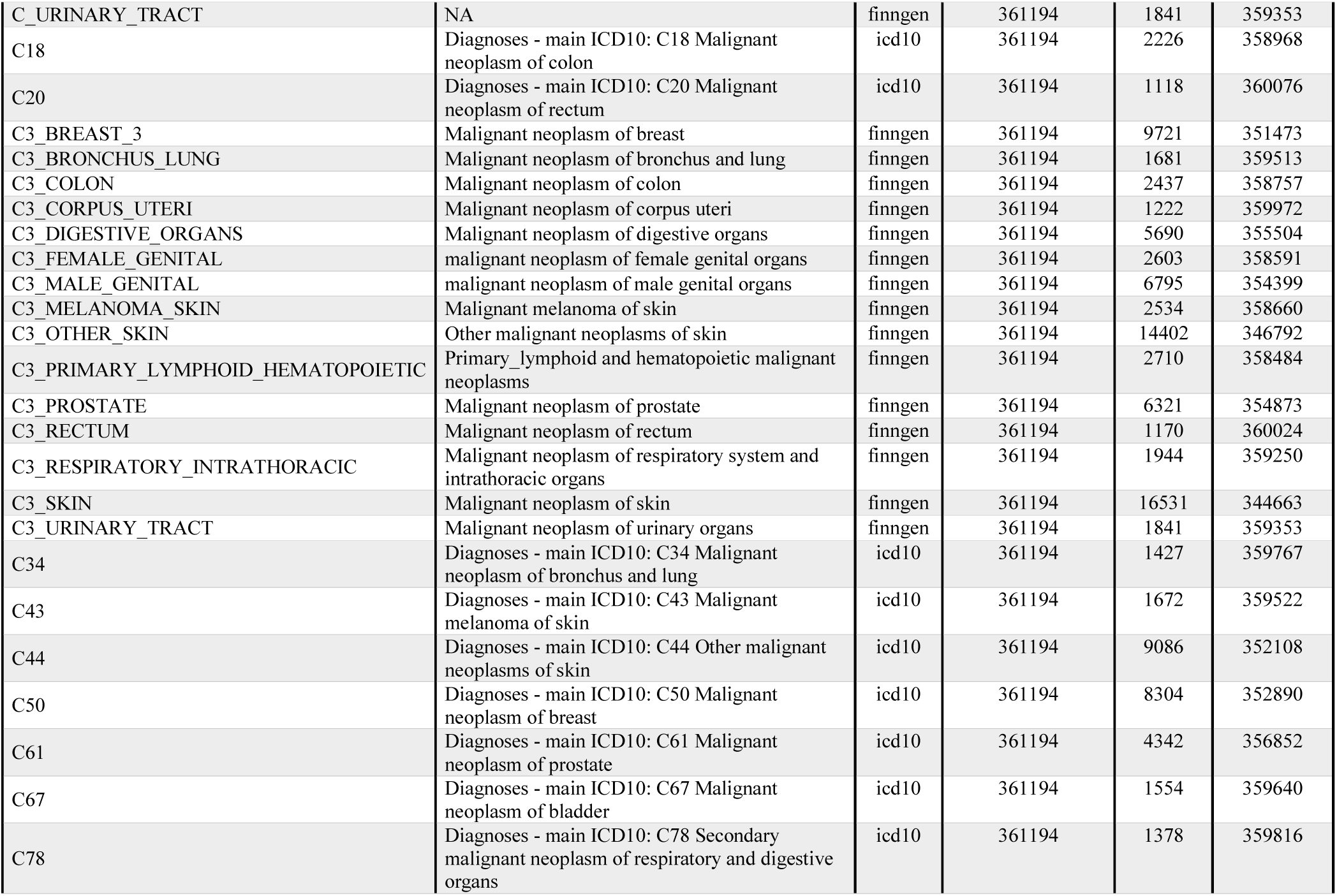
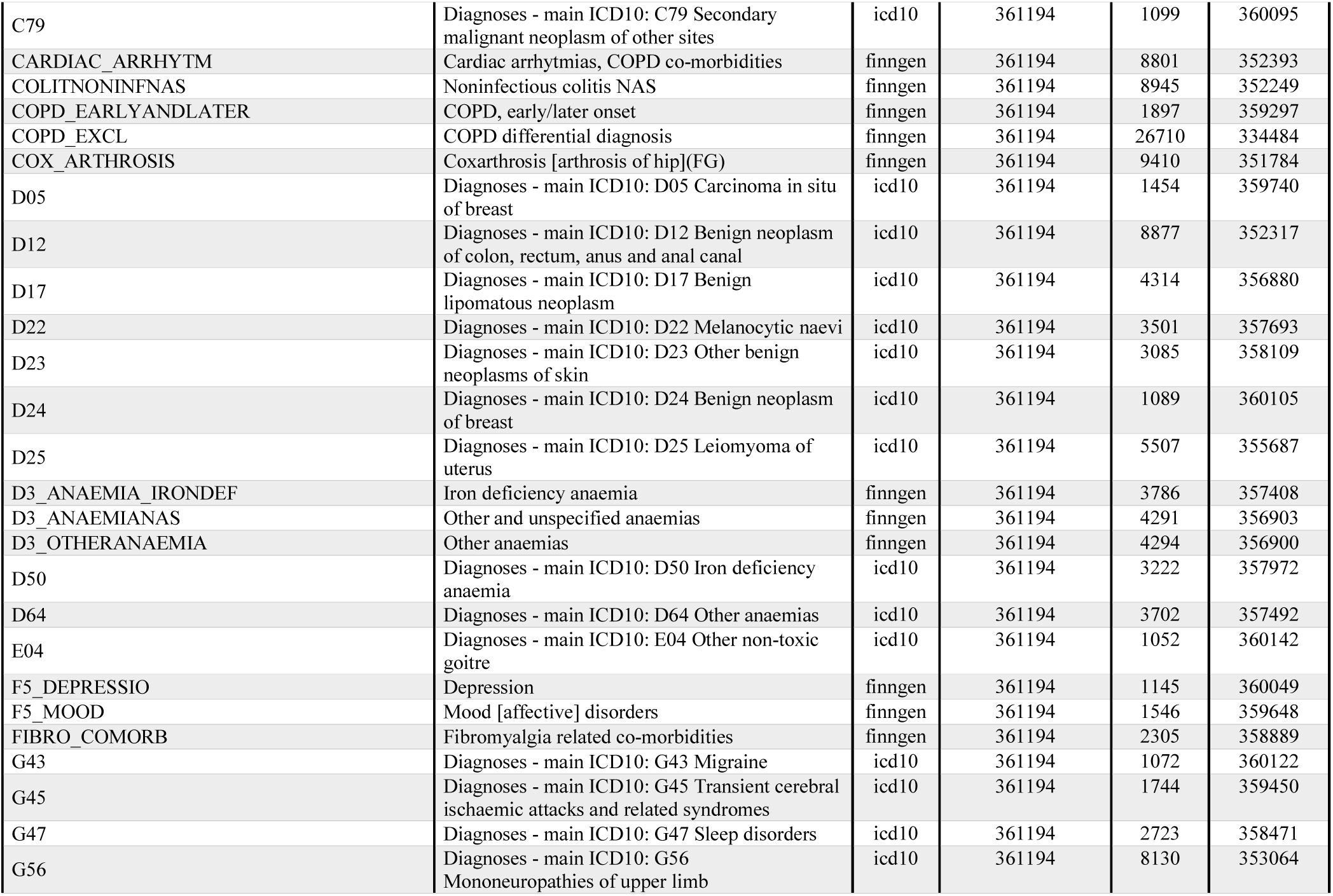
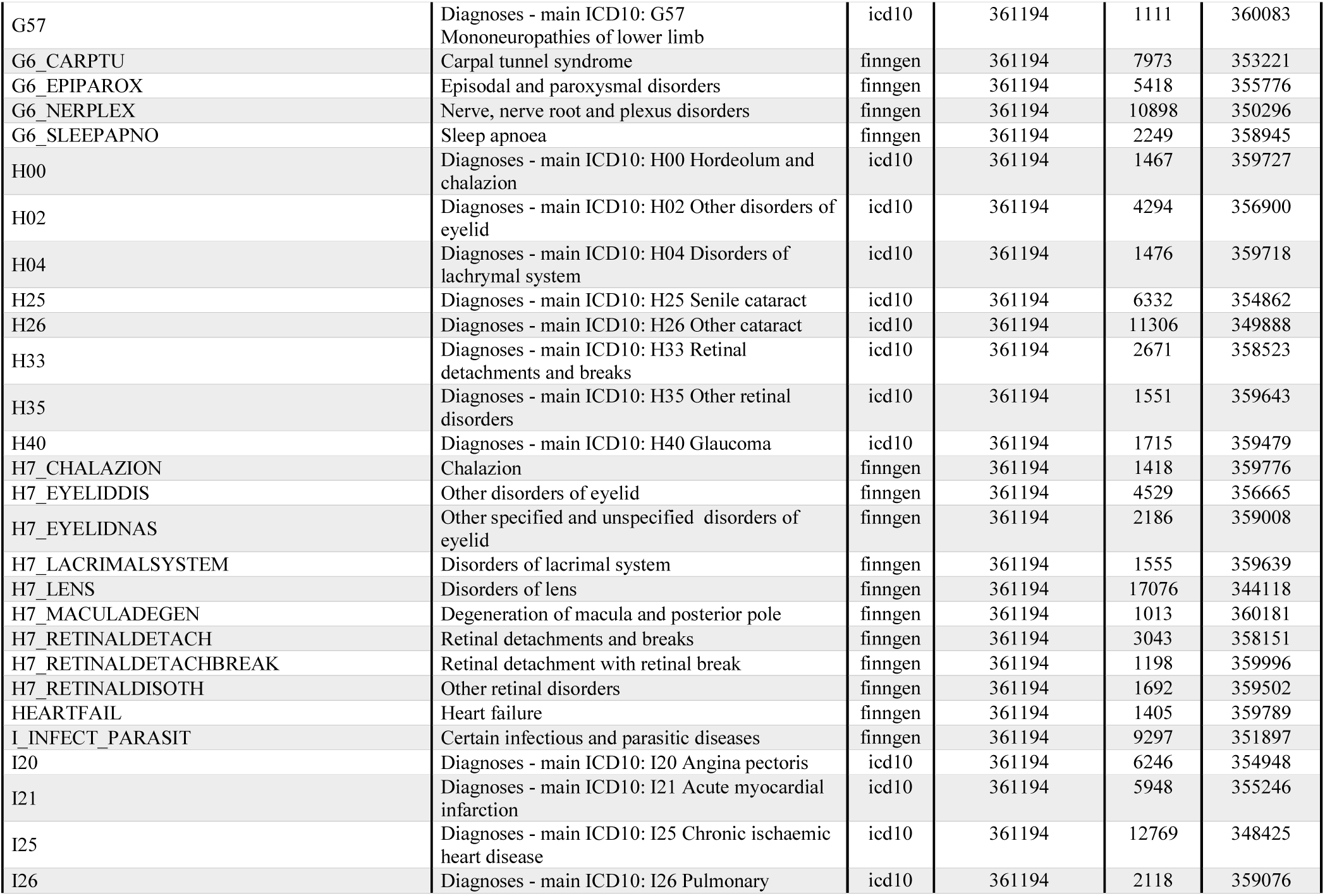
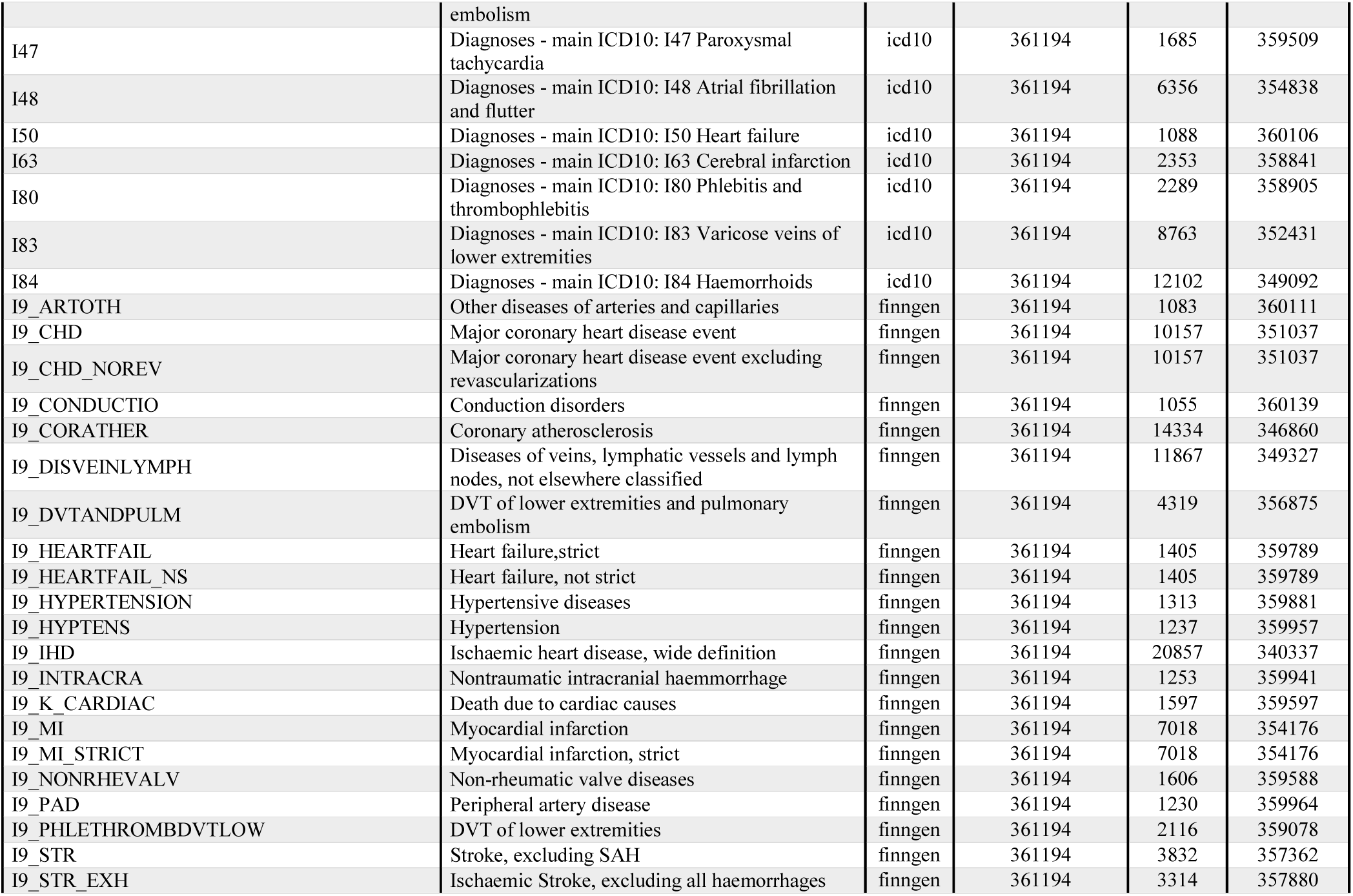
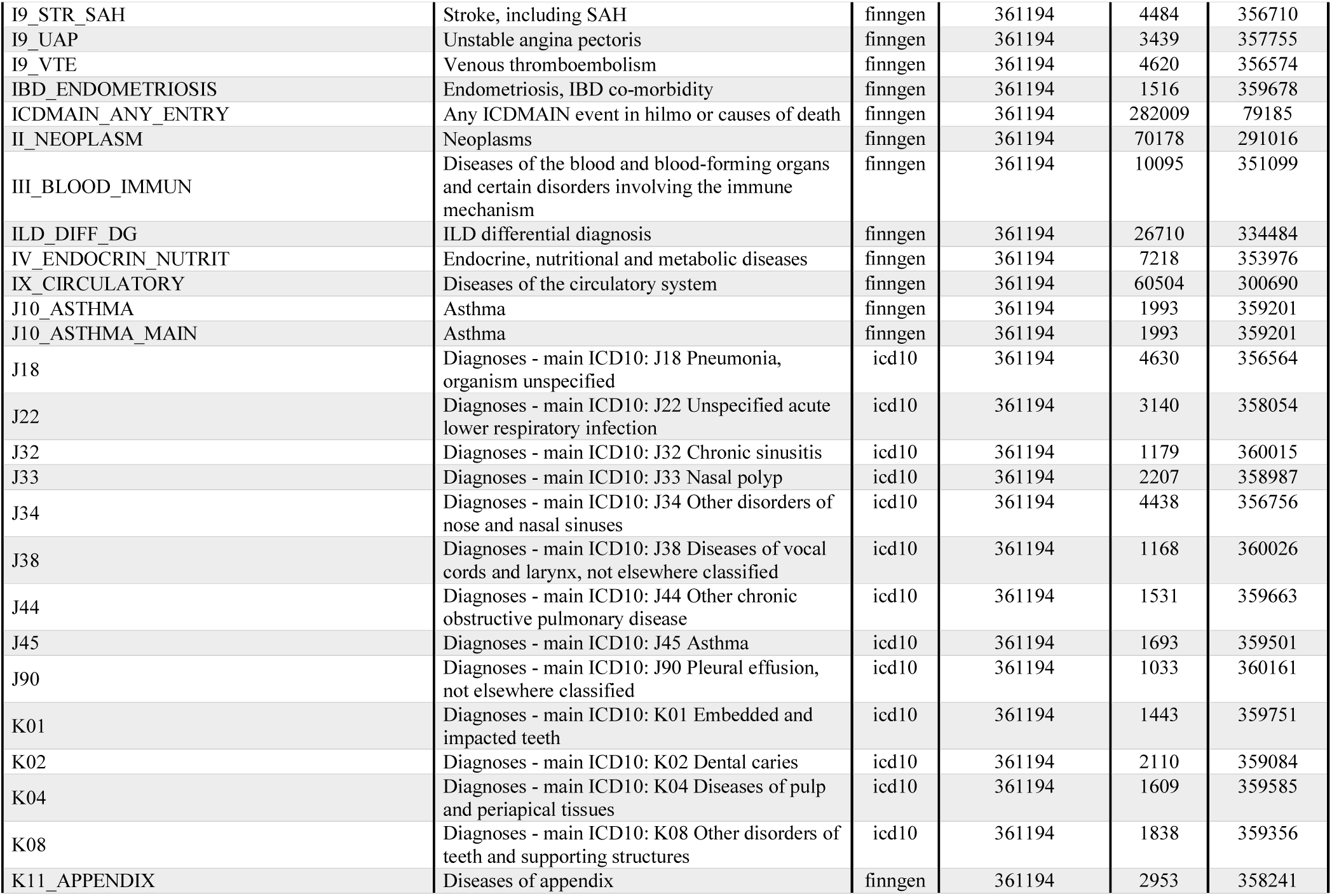
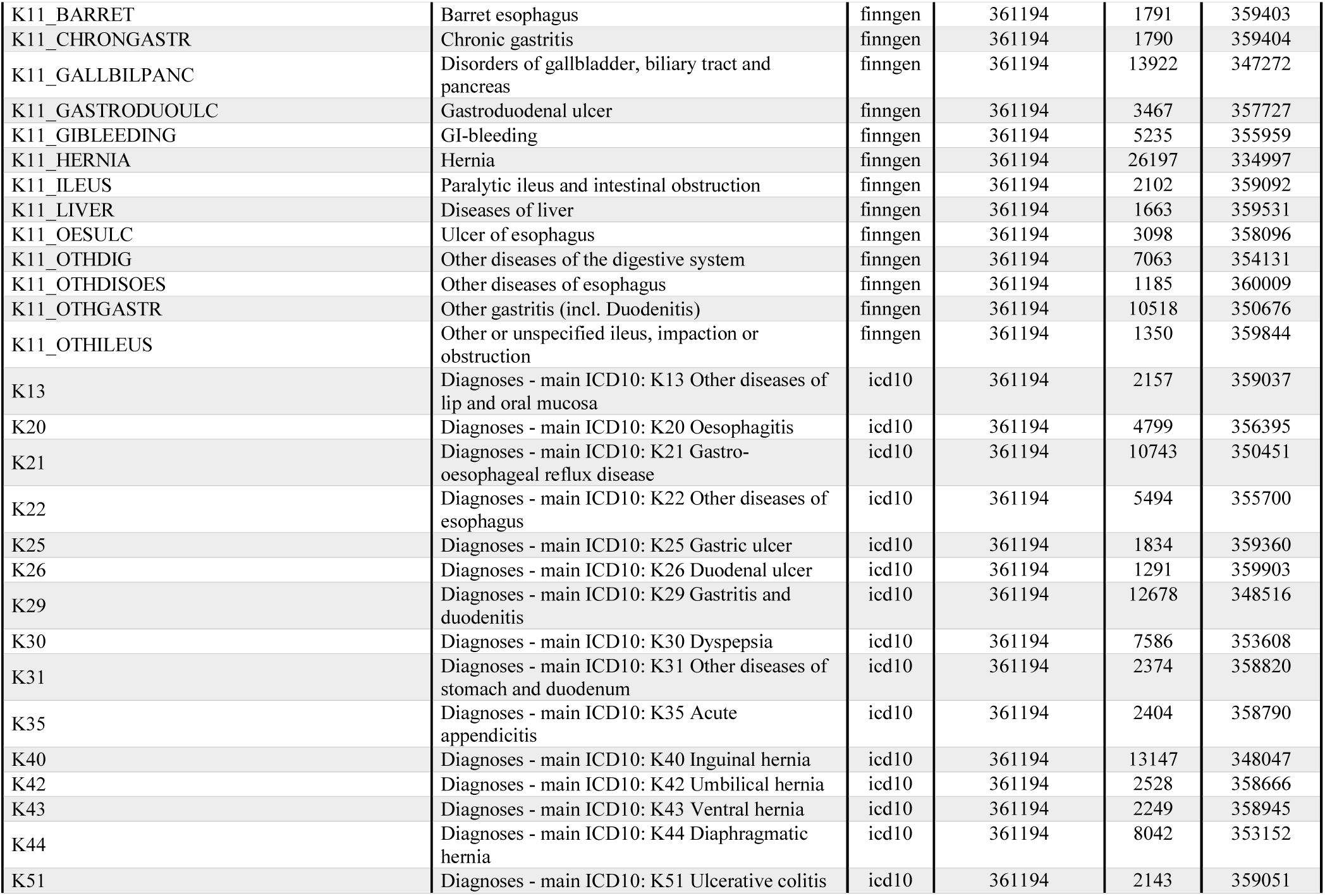
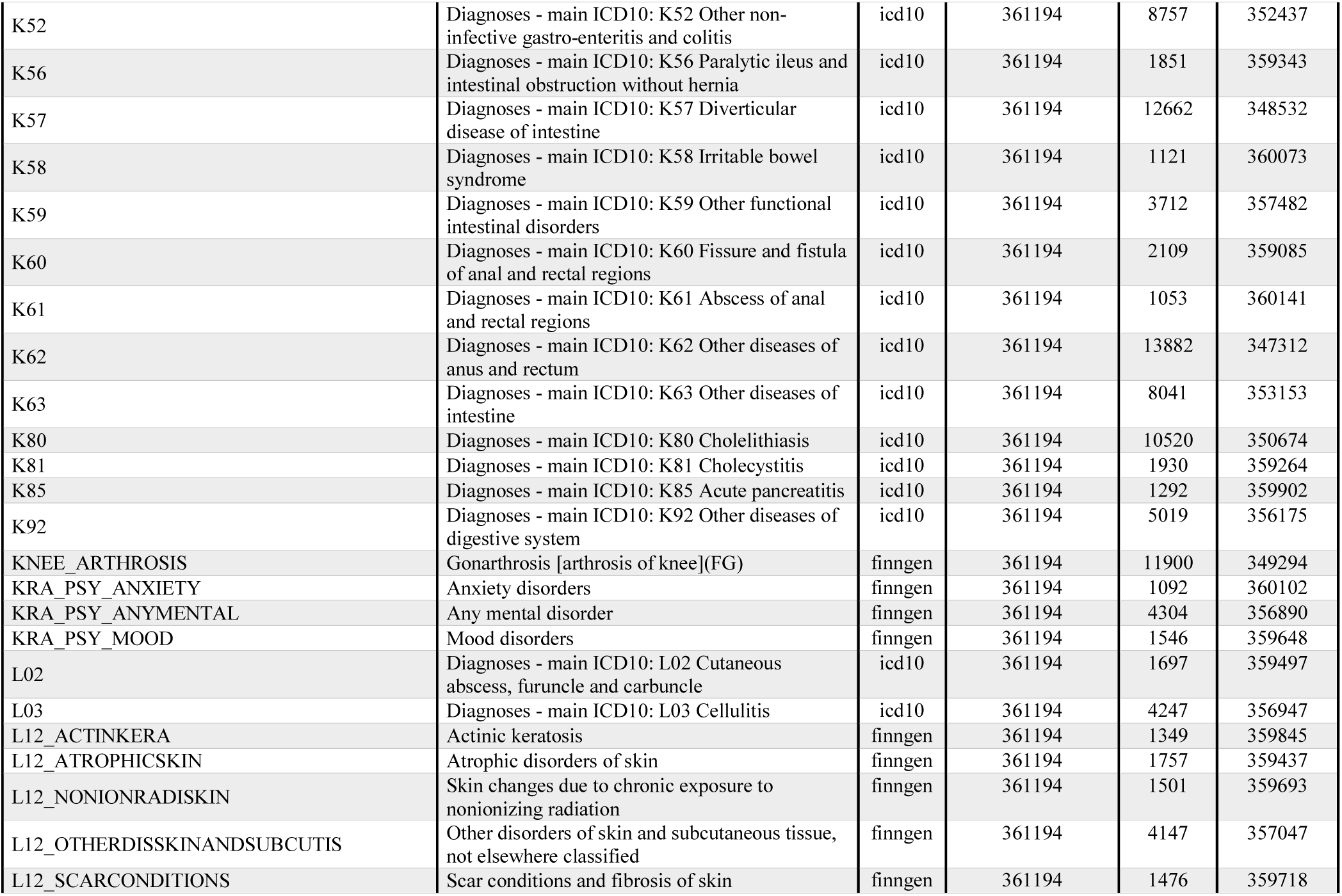
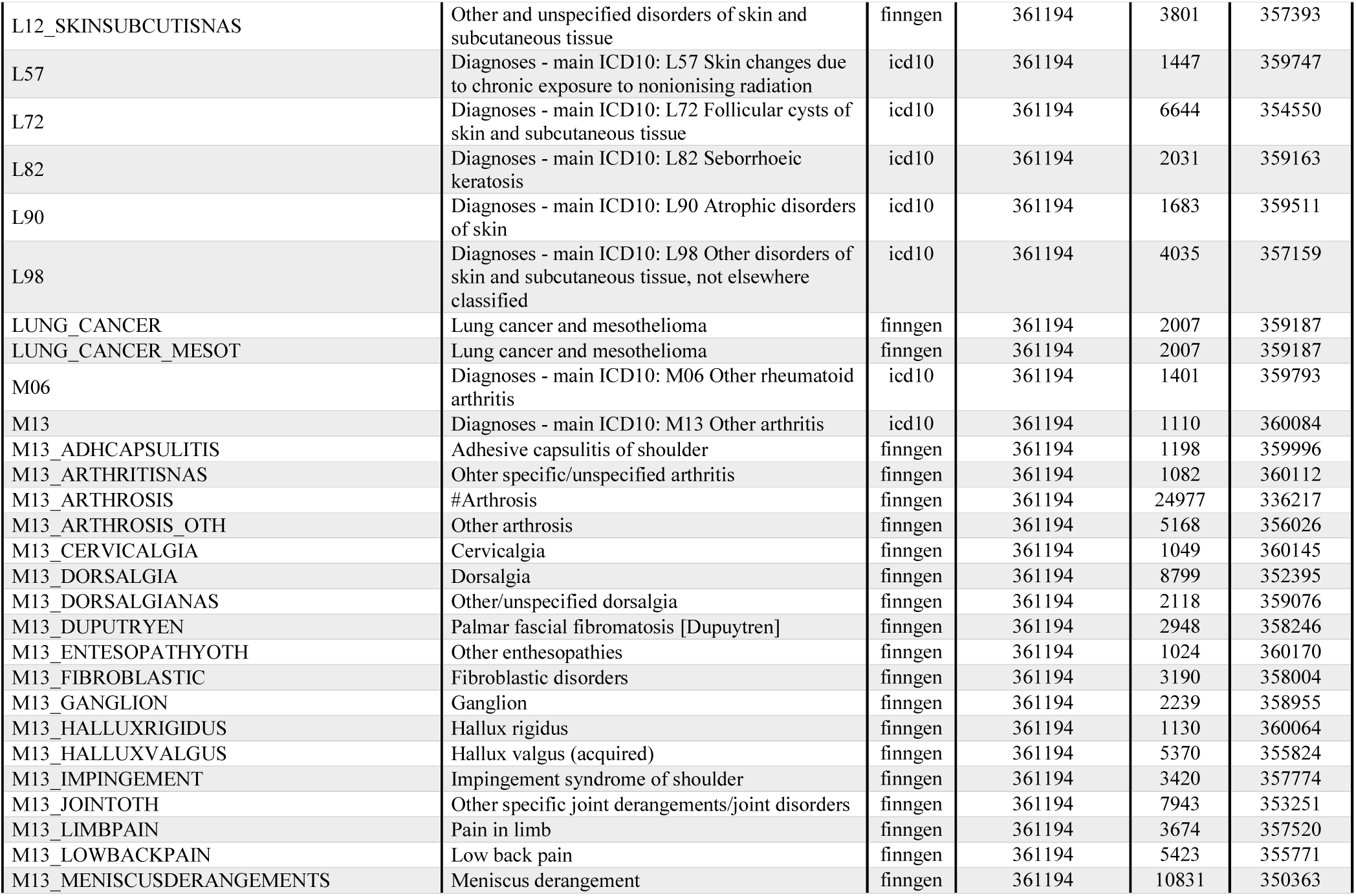
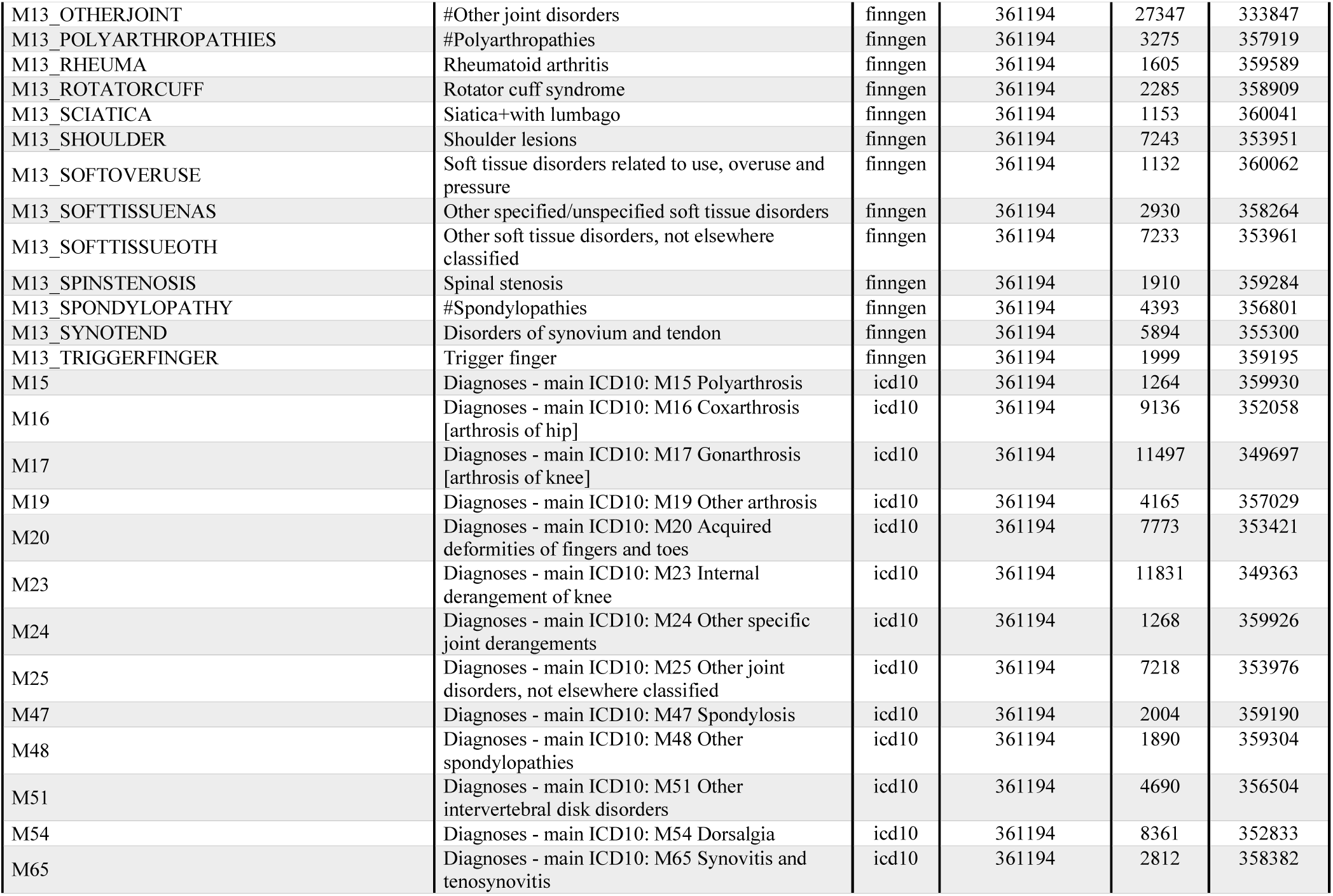
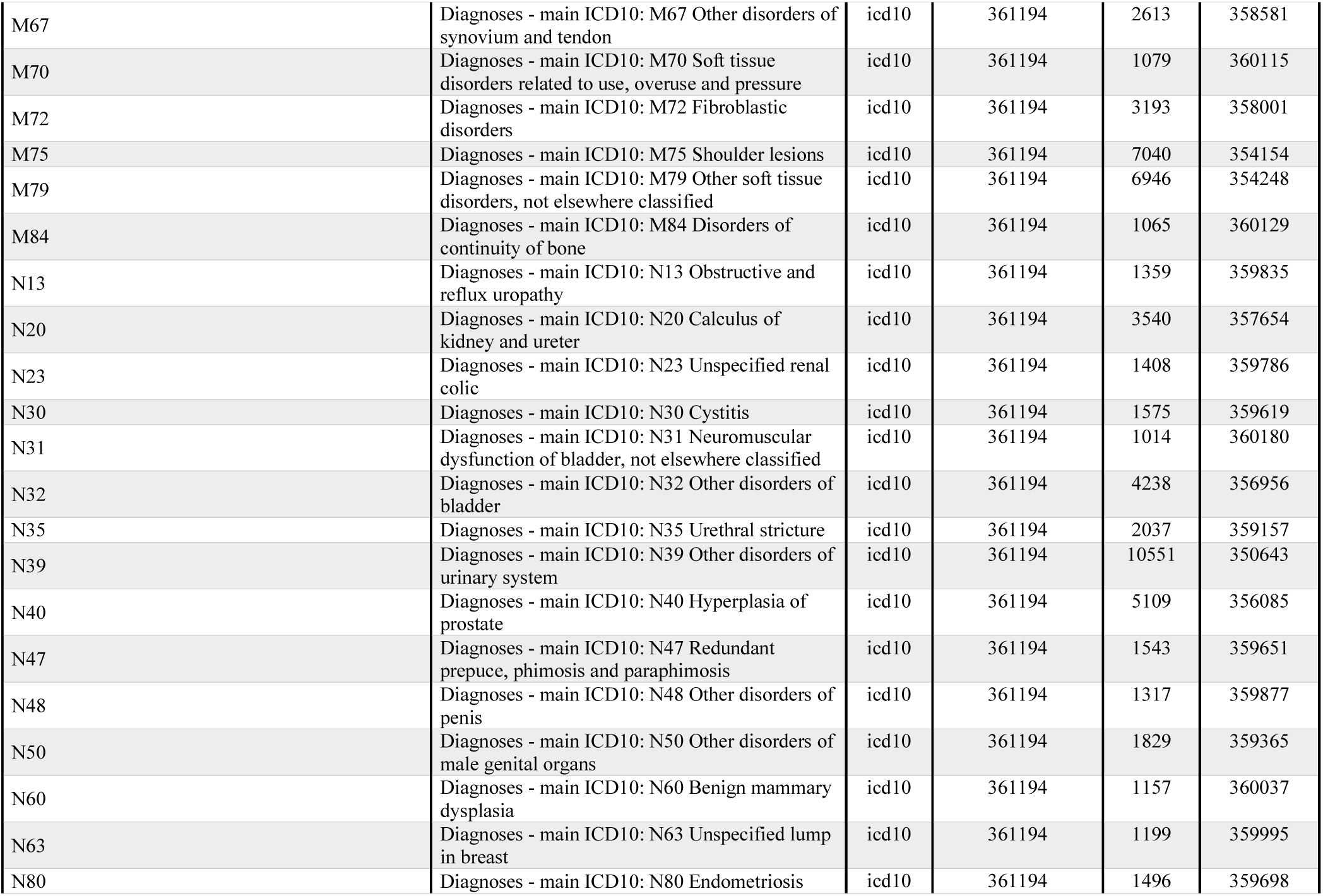
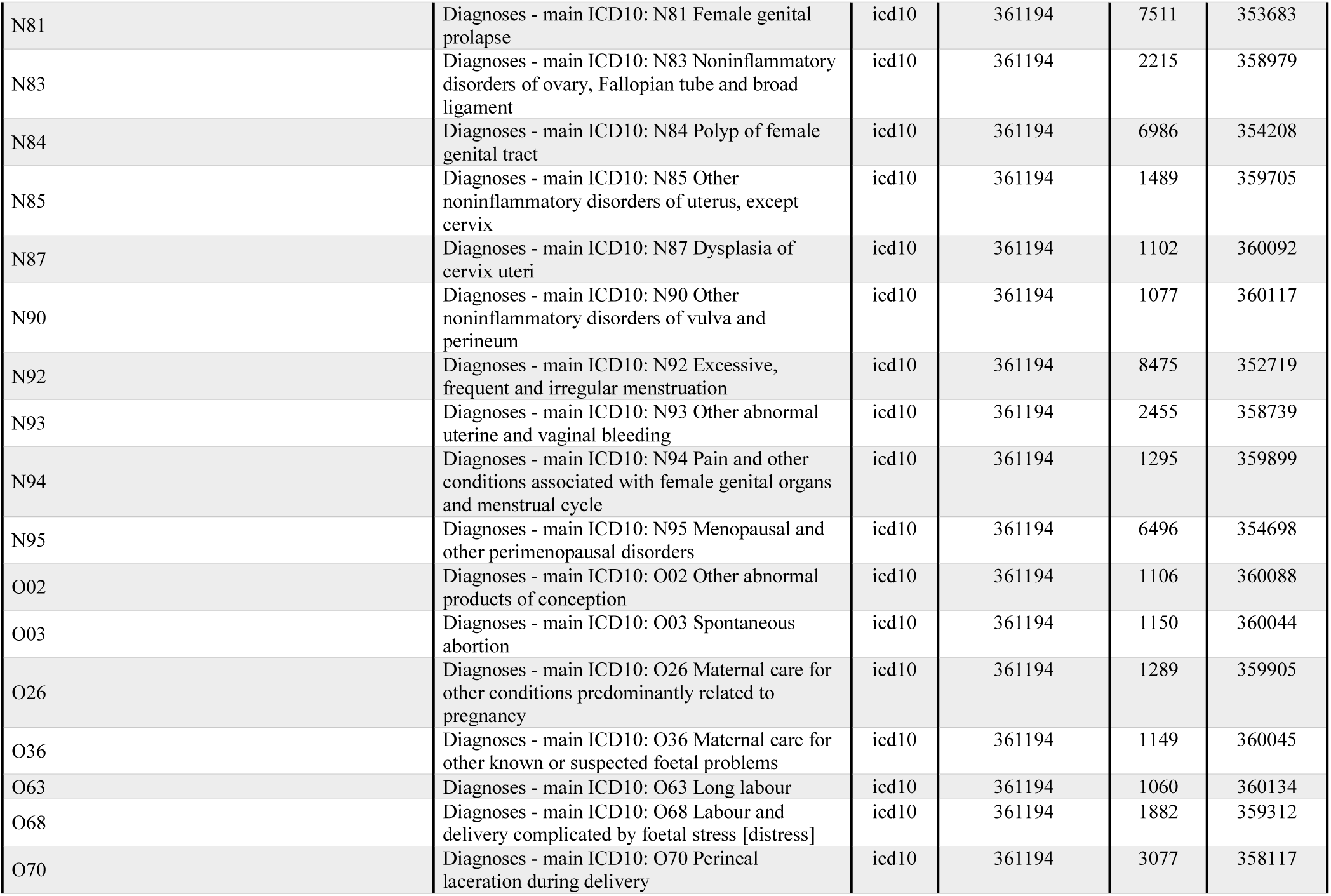
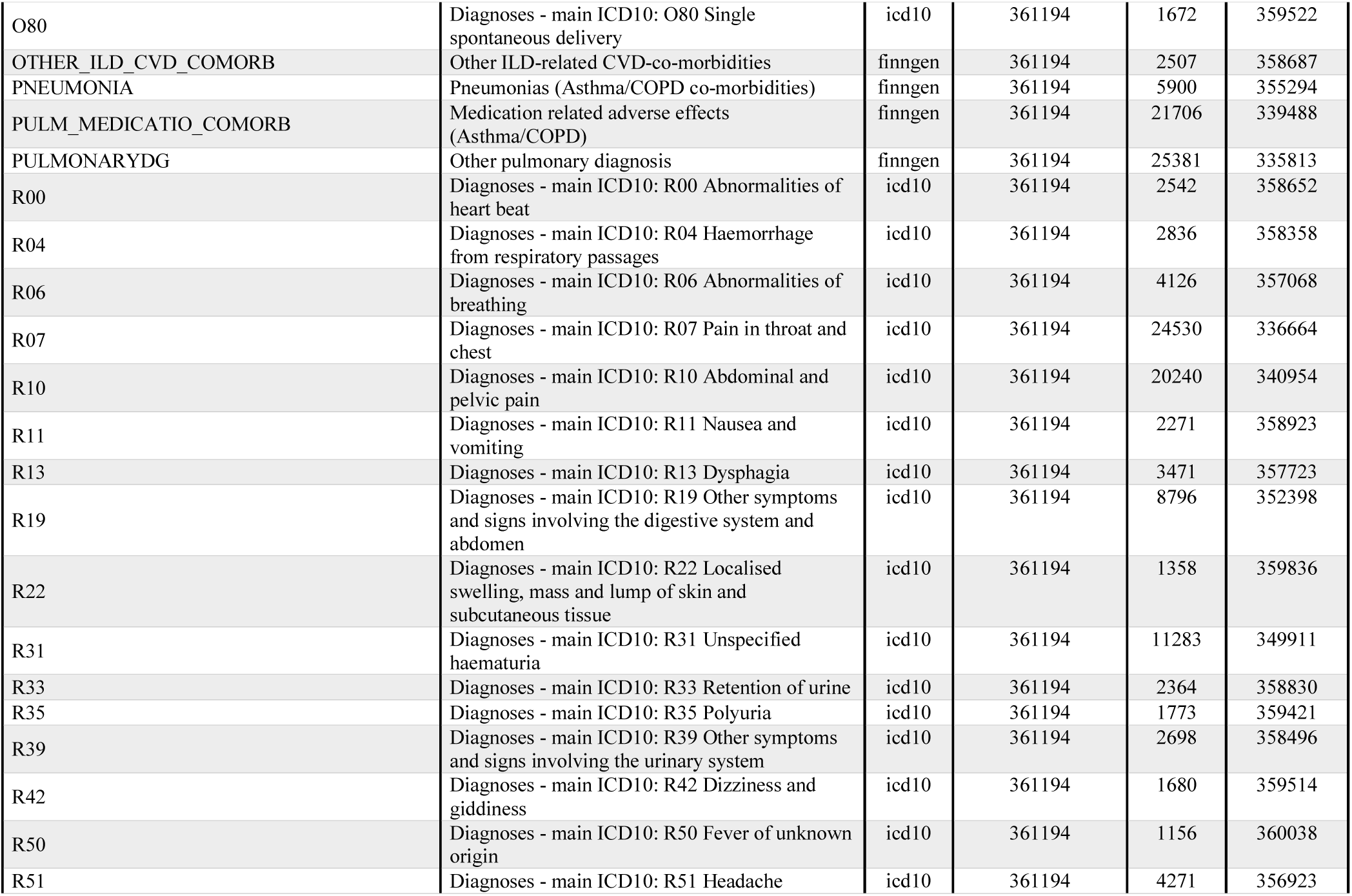
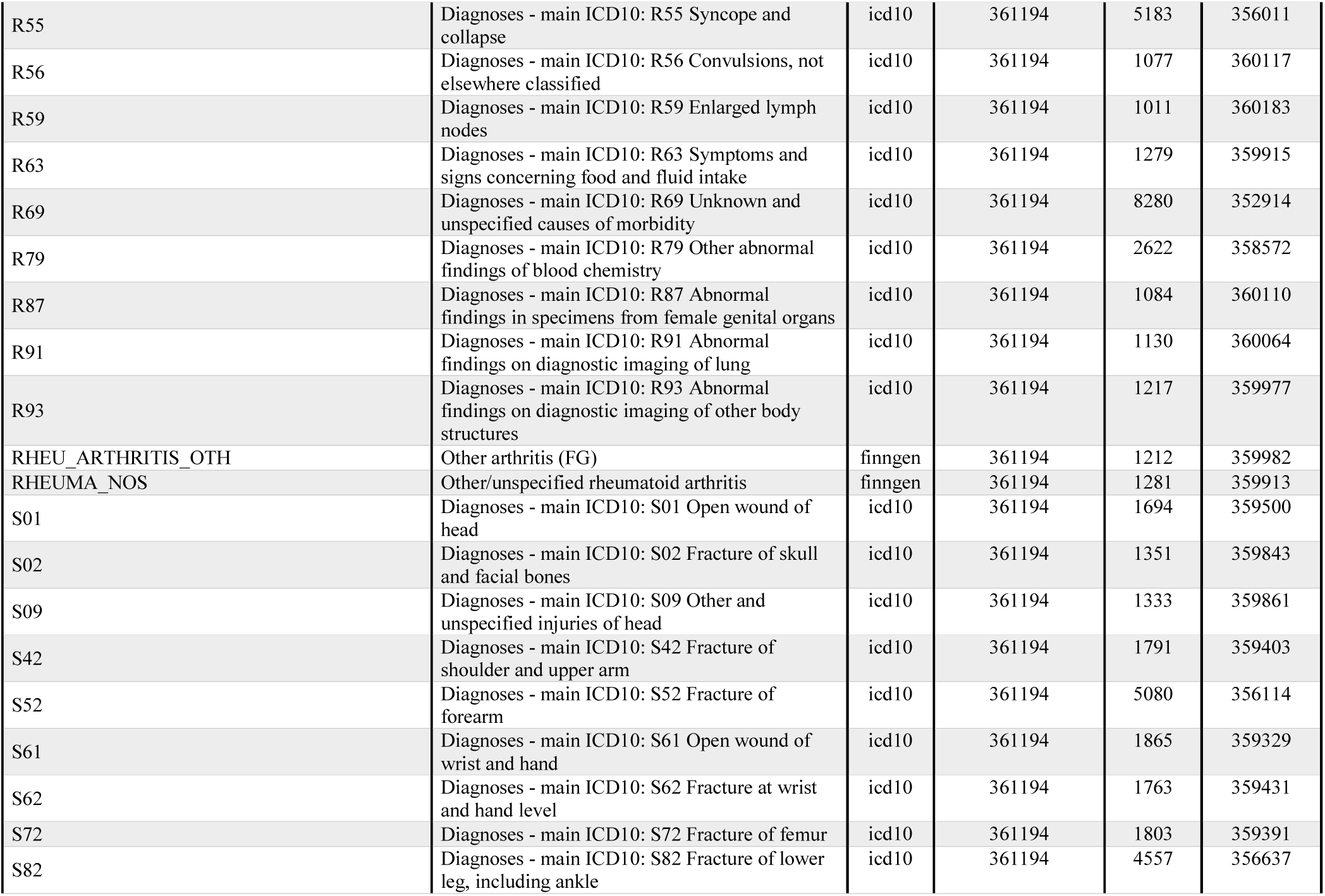
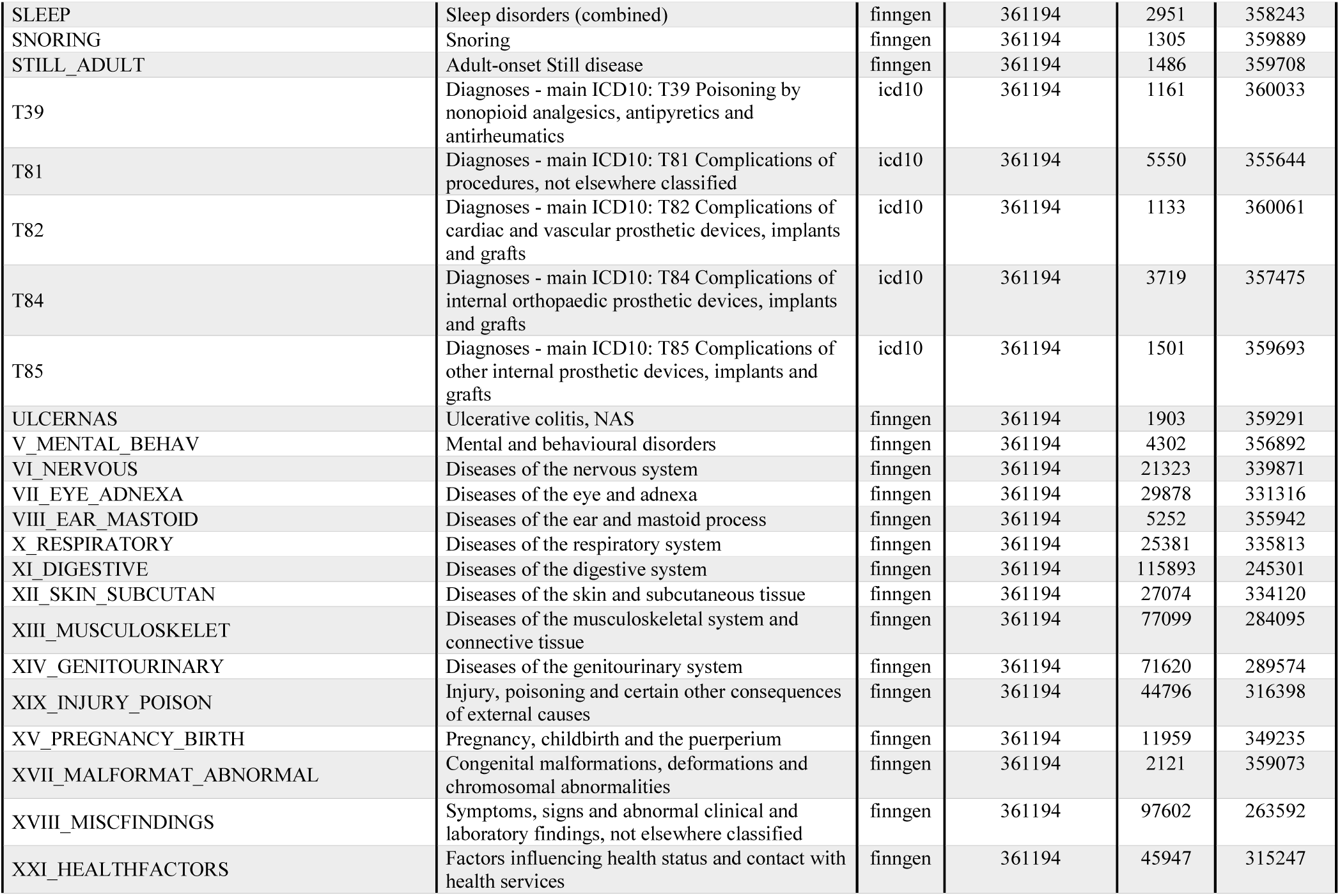
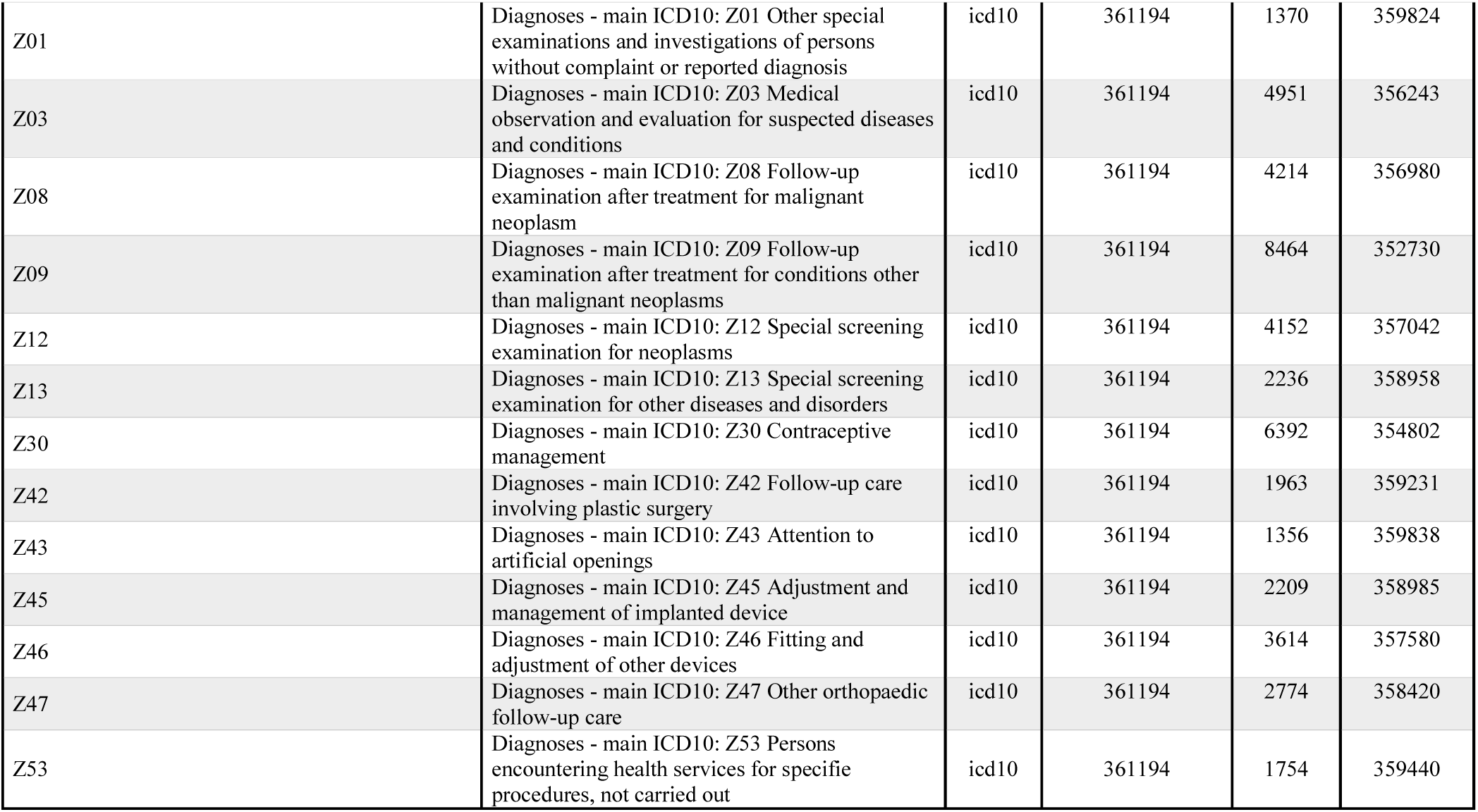
Clinically-relevant phenotypic traits investigated.

**Table S2:** Clinically-relevant phenotypes identified as significant in the *TTR* PheWAS. We report the Z scores observed in the overall analysis and in the sex-stratified analysis for both *TTR* and *RBP4*. Z scores surviving multiple testing correction are highlighted in bold. NA indicates associations not tested due the quality control applied. NS indicates phenotypes not significantly associated with *RBP4* variants.

| Traits | *TTR* rsid | Z Score |  |  | *RBP4* rsid | Z score |  |  |
| --- | --- | --- | --- | --- | --- | --- | --- | --- |
|  |  | Overall | Female | Male |  | Overall | Female | Male |
| I25 (Chronic ischemic heart disease) | rs140226130 | 2.41 | **4.75** | 0.06 | NS |  |  |  |
| I9_HEARTFAIL (Heart failure, strict) | rs73956431 | 4.27 | 0.65 | **4.69** | rs1326222 | **5.02** | **4.37** | 3.18 |
| I48 (Atrial fibrillation and flutter) | rs10163755 | 3.81 | 0.05 | **4.58** | rs4917692 | 1.54 | **4.39** | -1.15 |
| R13 (Dysphagia) | rs2949506 | 3.18 | **4.61** | -0.49 | rs61886346 | 2.52 | -0.35 | **4.31** |
| K63 (Other diseases of intestine) | rs970866 | -3.77 | -0.68 | **-4.49** | NS |  |  |  |
| N31 (Neuromuscular dysfunction of bladder, not elsewhere classified) | rs9953311 | **-4.73** | -3.58 | -3.07 | NS |  |  |  |
| N39 (Other disorders of urinary system) | rs71173870 | 3.59 | **4.42** | -0.14 | NS |  |  |  |
| N32 (Other disorders of bladder) | rs138038371 | 4.32 | 0.32 | **5.03** | NS |  |  |  |
| N20 (Calculus of kidney and ureter) | rs8090264 | 4.32 | 0.89 | **4.56** | NS |  |  |  |
| H26 (Other cataract) | rs12966815 | 3.02 | **4.40** | -0.33 | NS |  |  |  |
| H33 (Retinal detachments and breaks) | rs117637258 | 3.3 | **4.39** | 0.63 | NS |  |  |  |
| M13 (SPINSTENOSIS (Spinal stenosis) | rs116937108 | **5.29** | 4.24 | 3.24 | rs112288944 | 1.87 | -1.61 | **4.34** |
| KRA_PSY_ANXIETY (Anxiety disorders) | rs554521234 | **4.44** | 4.8 | 1.14 | rs112059561 | **4.51** | 2.99 | 3.49 |
| M13_FIBROBLASTIC (Fibroblastic disorders) | rs60487427 | -3.6 | 0.49 | **-4.46** | NS |  |  |  |
| M13_SOFTTISSUES (Other specified/unspecified soft tissue disorders) | rs558461933 | **4.68** | **4.42** | 2.11 | NS |  |  |  |
| M75 (Shoulder lesions) | rs77728273 | -3.34 | **-4.38** | -0.19 | NS |  |  |  |
| K61 (Abscess of al and rectal regions) | rs72948149 | **4.77** | 2.72 | 3.91 | NS |  |  |  |
| K60 (Fissure and fistula of al and rectal regions) | rs72927361 | **-4.76** | -3.39 | -3.38 | NS |  |  |  |
| K11_BARRET (Barret esophagus) | rs147570462 | **5.30** | 2.56 | **4.64** | rs12573026 | **5.33** | 1.72 | **5.33** |
| K11_OESULC (Ulcer of esophagus) | rs74481699 | **4.51** | 1.82 | 4.29 | NS |  |  |  |
| C18 (Malign neoplasm of colon) | rs78431500 | 2.07 | -2.14 | **4.68** | rs142083973 | 3.7 | 0.33 | **4.65** |
| C3_PRIMARY_LYMPHOID_HEMATOPOIETIC (Primary lymphoid and hematopoietic malign neoplasms) | rs17718949 | **4.51** | 3.63 | 2.81 | NS |  |  |  |
| T85 (Complications of other internal prosthetic devices, implants and grafts) | rs34444556 | **4.54** | NA | 4.20 | NS |  |  |  |
| T81 (Complications of procedures, not elsewhere classified) | rs182526571 | 2.4 | -0.83 | **4.51** | NS |  |  |  |
| Z45 (Adjustment and management of implanted device) | rs117672477 | **4.51** | 2.93 | 3.43 | NS |  |  |  |
| O02 (Other abnormal products of conception) | rs139327590 | 4.40 | **4.38** | NA | NS |  |  |  |

**Table S3:**
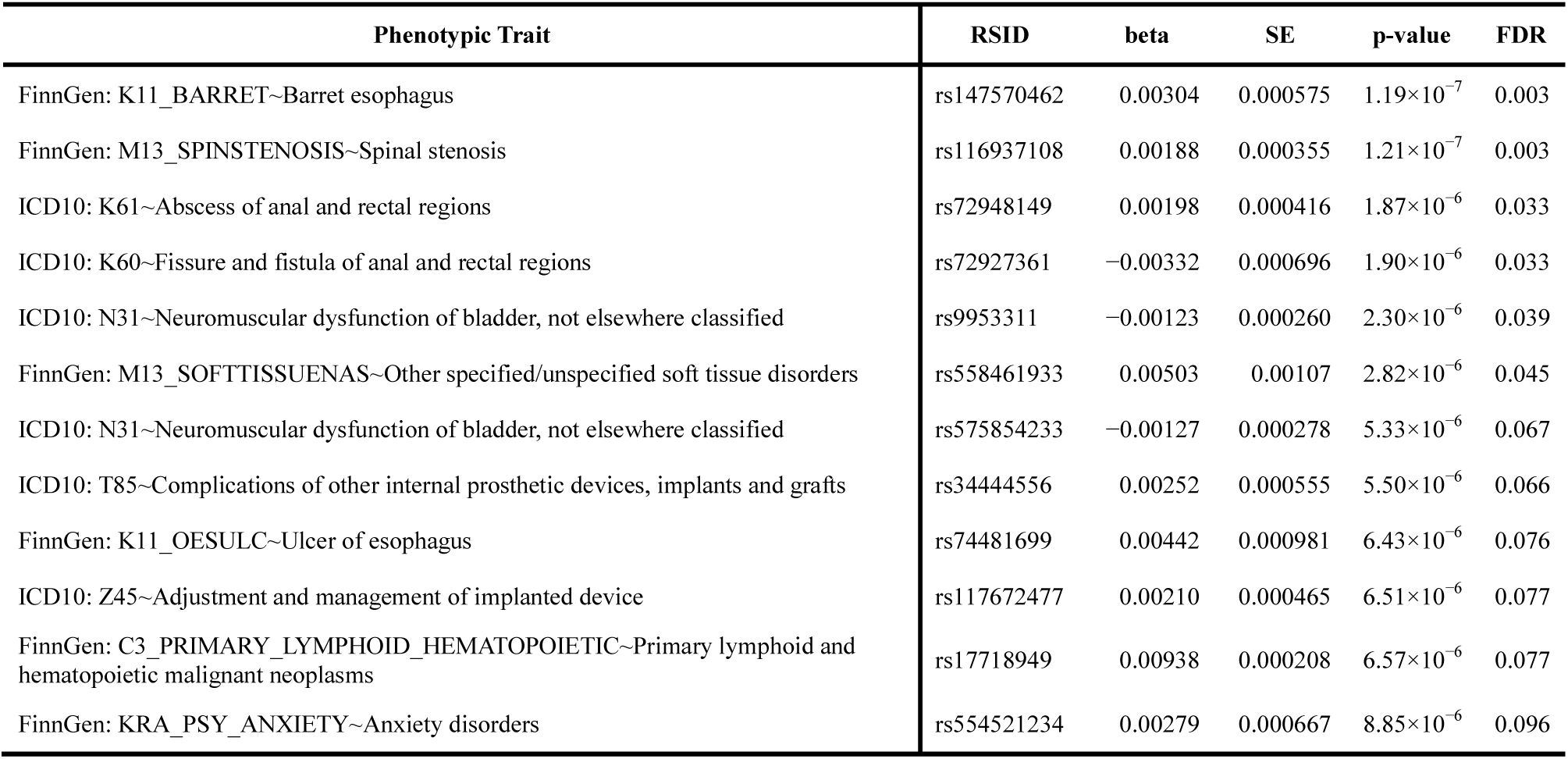
Variants localized on *TTR* target genomic region (NC_000018.9: 27,171,000 – 31,171,500) and their significantly associations with phenotypic traits related in the overall sample. Phenotypic traits are reported with respectively ICD-10 and FinnGen codes. Information about beta value, SE, p-value and FDR q-value are also reported. Information about allele frequency and minor allele frequency are reported in Table S10.

**Table S4:** Variants localized on *TTR* target genomic region (NC_000018.9: 27,171,000 – 31,171,500) and their significantly associations with phenotypic traits related in the female-specific analysis. Phenotypic traits are reported with respectively ICD-10 and FinnGen codes. Information about beta value, SE, p-value and FDR q-value are also reported. Information about allele frequency and minor allele frequency are reported in Table S10.

| Phenotypic Trait | RSID | beta | SE | p-value | FDR |
| --- | --- | --- | --- | --- | --- |
| ICD10: I25~Chronic ischaemic heart disease | rs140226130 | 0.00338 | 0.000711 | $2.00 \times 10^{-6}$ | 0.029 |
| ICD10: R13~Dysphagia | rs2949506 | 0.00195 | 0.000422 | $3.95 \times 10^{-6}$ | 0.047 |
| ICD10: N39~Other disorders of urinary system | rs71173870 | 0.00298 | 0.000675 | $9.81 \times 10^{-6}$ | 0.079 |
| FinnGen: M13_SOFTTISSUENAS~Other specified/unspecified soft tissue disorders | rs558461933 | 0.00646 | 0.00146 | $9.92 \times 10^{-6}$ | 0.079 |
| ICD10: H26~Other cataract | rs12966815 | 0.00253 | 0.000576 | $1.10 \times 10^{-5}$ | 0.079 |
| ICD10: H33~Retinal detachments and breaks | rs117637258 | 0.00290 | 0.000660 | $1.12 \times 10^{-5}$ | 0.079 |
| ICD10: O02~Other abnormal products of conception | rs139327590 | 0.00389 | 0.000888 | $1.16 \times 10^{-5}$ | 0.079 |
| ICD10: M75~Shoulder lesions | rs77728273 | -0.00379 | 0.000866 | $1.21 \times 10^{-5}$ | 0.079 |

**Table S5:** Variants localized on *TTR* target genomic region (NC_000018.9: 27,171,000 – 31,171,500) and their significantly associations with phenotypic traits related in the male-specific analysis. Phenotypic traits are reported with respectively ICD-10 and FinnGen codes. Information about beta value, SE, p-value and FDR q-value are also reported. Information about allele frequency and minor allele frequency are reported in Table S10.

| Phenotypic Trait | RSID | beta | SE | p-value | FDR |
| --- | --- | --- | --- | --- | --- |
| ICD10: N32~Other disorders of bladder | rs138038371 | 0.00893 | 0.00178 | $5.01 \times 10^{-7}$ | 0.010 |
| FinnGen: I9_HEARTFAIL~Heart failure, strict | rs73956431 | 0.00529 | 0.00113 | $2.74 \times 10^{-6}$ | 0.042 |
| ICD10: C18~Malignant neoplasm of colon | rs78431500 | 0.00579 | 0.00124 | $2.89 \times 10^{-6}$ | 0.044 |
| FinnGen: K11_BARRET~Barret esophagus | rs147570462 | 0.00466 | 0.001 | $3.45 \times 10^{-6}$ | 0.051 |
| ICD10: I48~Atrial fibrillation and flutter | rs10163755 | 0.00299 | 0.000653 | $4.63 \times 10^{-6}$ | 0.064 |
| ICD10: N20~Calculus of kidney and ureter | rs8090264 | 0.00208 | 0.000455 | $5.23 \times 10^{-6}$ | 0.071 |
| ICD10: T81~Complications of procedures, not elsewhere classified | rs182526571 | 0.00752 | 0.00167 | $6.57 \times 10^{-6}$ | 0.083 |
| ICD10: K63~Other diseases of intestine | rs970866 | -0.00507 | 0.00113 | $7.14 \times 10^{-6}$ | 0.086 |
| FinnGen: M13_FIBROBLASTIC~Fibroblastic disorders | rs60487427 | -0.00373 | 0.000838 | $8.38 \times 10^{-6}$ | 0.096 |

**Table S6:** *TTR* coding mutations in the UK Biobank sample investigated. Rsid, localization, imputation score, allelic frequency (AF), precursor and/or protein substitution, consequences of variants are reported.

| RSID | variant | info_score | AF | Precursor_TTR | TTR | Consequences |
| --- | --- | --- | --- | --- | --- | --- |
| rs138657343 | 18:29171879:G:A | 0.823 | 0.0003 | Arg5His | - | Missense_variant |
| rs1800458 | 18:29172865:G:A | 1 | 0.0814 | Gly26Ser | Gly26Ser | Missense_variant |
| rs121918074 | 18:29175210:C:A | 0.863 | 0.00005 | His110Asn | His90Asn | Missense_variant |
| rs28933981 | 18:29178610:C:T | 0.925 | 0.0037 | Thr139Met | Thr119Met | Missense_variant |
| rs2276382 | 18:29178611:G:A | 0.823 | 0.00005 | Thr139= | Thr119= | Synonymous_variant |
| rs76992529 | 18:29178618:G:A | 1 | 0.00008 | Val142Ile | Val122Ile | Missense_variant |

**Table S7:** Variants localized on *RBP4* target genomic region (NC_000010.10: 93,353,000 – 97,353,500) and their significantly associations with phenotypic traits in the overall-sample analysis. Phenotypic traits are reported with respectively ICD-10 and FinnGen codes. Information about beta value, SE, p-value and FDR q-value are also reported. Information about allele frequency and minor allele frequency are reported in Table S10.

| Phenotypic Trait | RSID | beta | SE | p-value | FDR |
| --- | --- | --- | --- | --- | --- |
| FinnGen: K11_BARRET~Barret esophagus | rs12573026 | 0.00124 | 0.000232 | $9.96\times 10^{-8}$ | 0.002 |
| FinnGen: I9_HEARTFAIL~Heart failure, strict | rs1326222 | 0.000935 | 0.000186 | $5.25\times 10^{-7}$ | 0.006 |
| FinnGen: KRA_PSY_ANXIETY~Anxiety disorders | rs112059561 | 0.00228 | 0.000506 | $6.44\times 10^{-6}$ | 0.042 |

**Table S8:** Variants localized on *RBP4* target genomic region (NC_000010.10: 93,353,000 – 97,353,500) and their significantly associations with phenotypic traits in the female-specific analysis. Phenotypic traits are reported with respectively ICD-10 and FinnGen codes. Information about beta value, SE, p-value and FDR q-value are also reported. Information about allele frequency and minor allele frequency are reported in Table S10.

| Phenotypic Trait | RSID | beta | SE | p-value | FDR |
| --- | --- | --- | --- | --- | --- |
| ICD10: I48~Atrial fibrillation and flutter | rs4917692 | 0.00152 | 0.000346 | $1.15\times 10^{-5}$ | 0.076 |
| FinnGen: I9_HEARTFAIL~Heart failure, strict | rs1326222 | 0.000802 | 0.000183 | $1.23\times 10^{-5}$ | 0.076 |

**Table S9:**
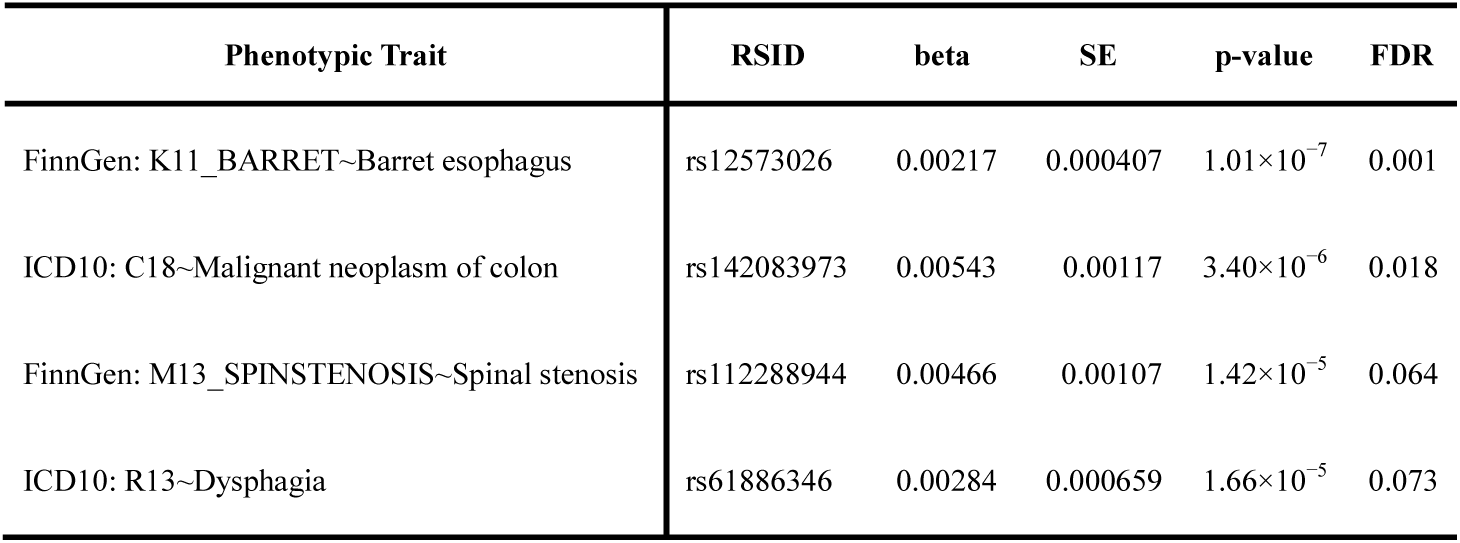
Variants localized on *RBP4* target genomic region (NC_000010.10: 93,353,000 – 97,353,500) and their significantly associations with phenotypic traits in the male-specific analysis. Phenotypic traits are reported with respectively ICD-10 and FinnGen codes. Information about beta value, SE, p-value and FDR q-value are also reported. Information about allele frequency and minor allele frequency are reported in Table S10.

**Table S10:**
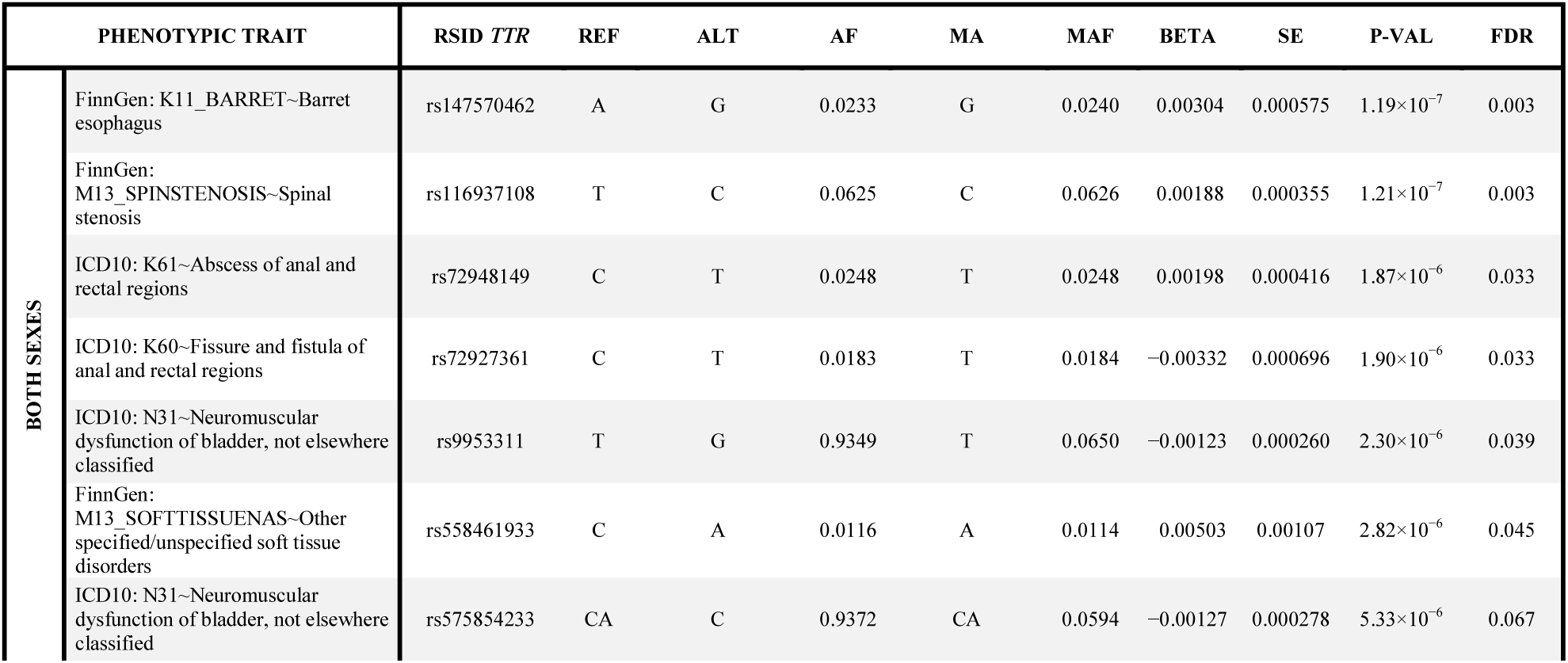
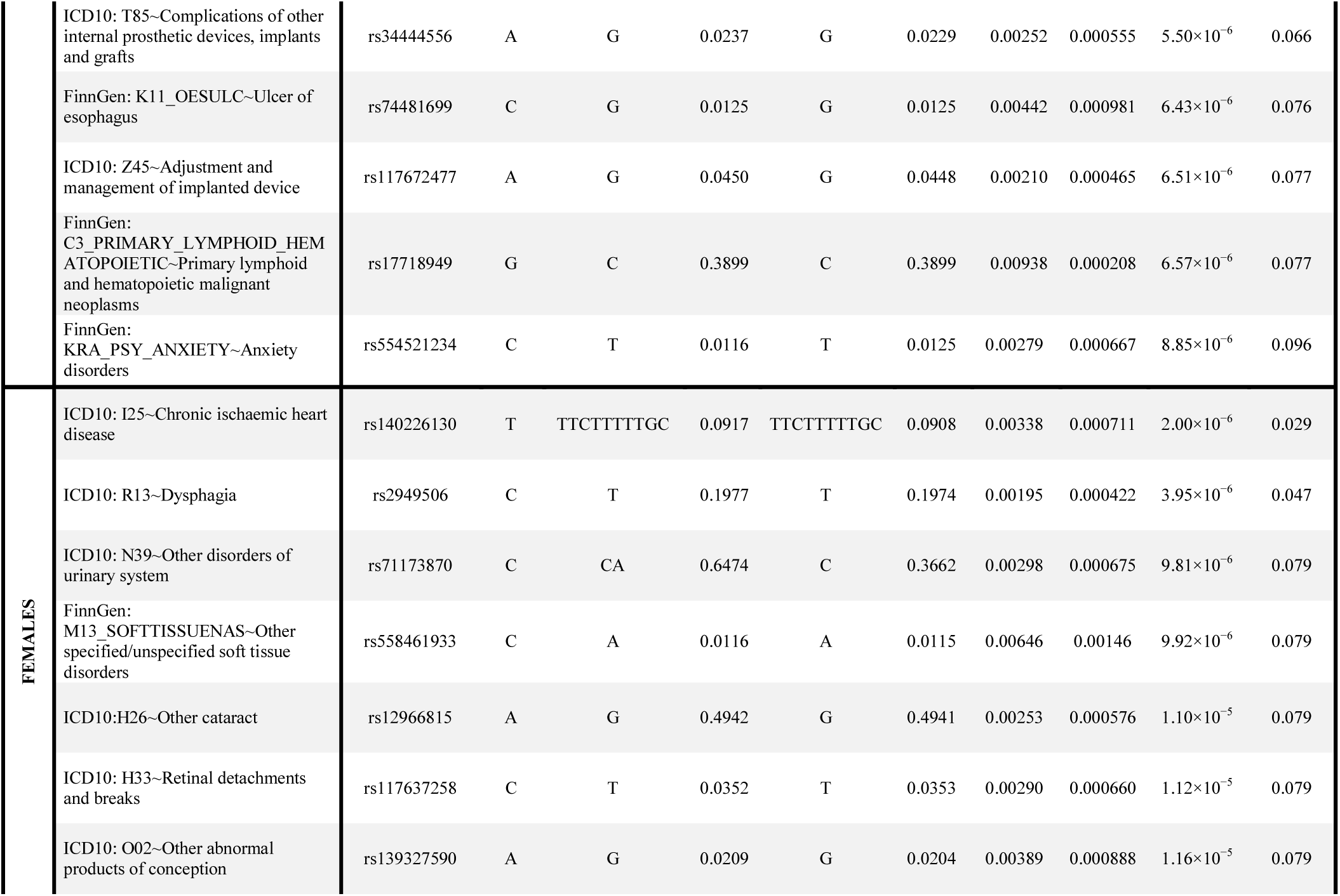
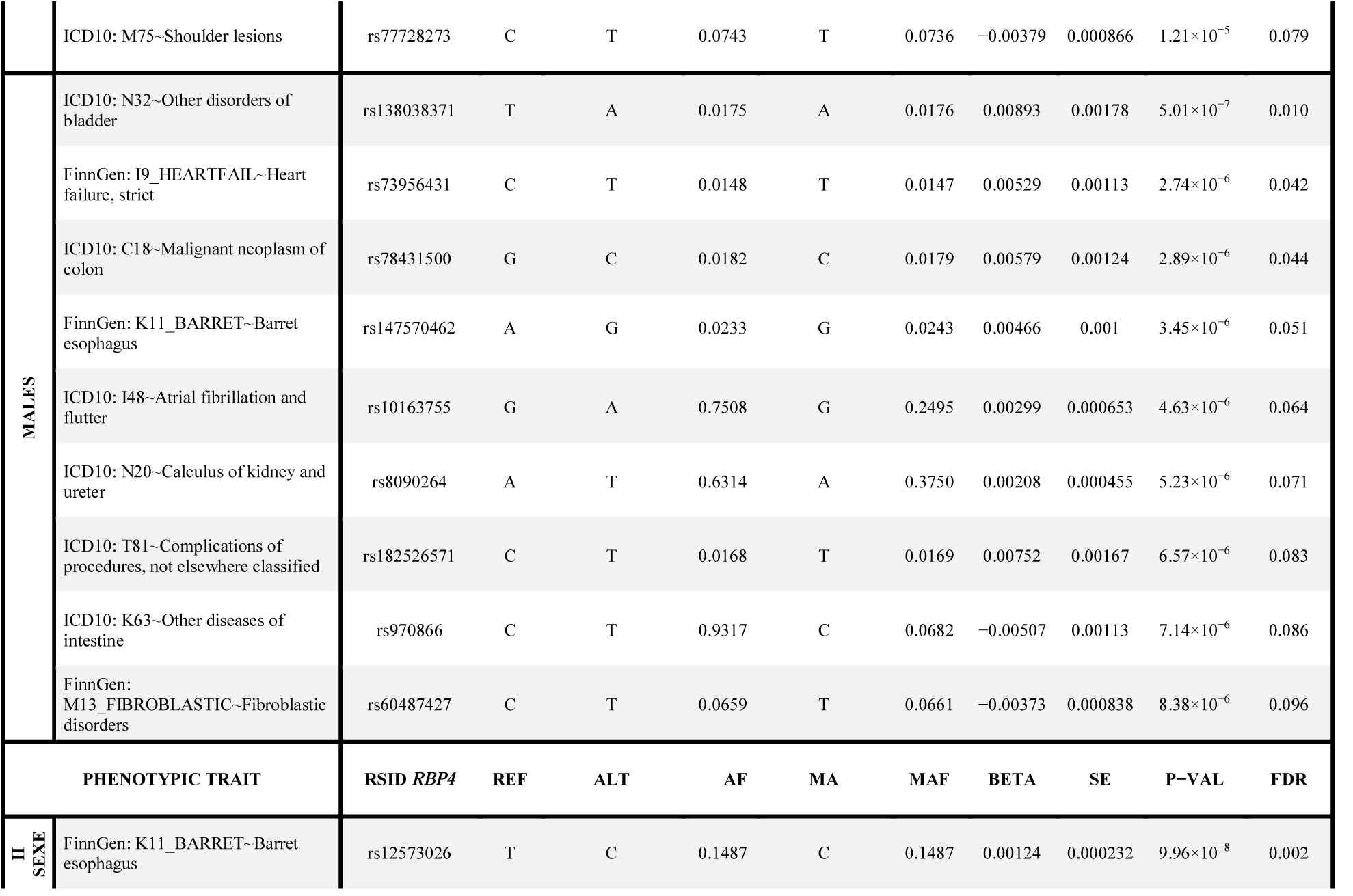
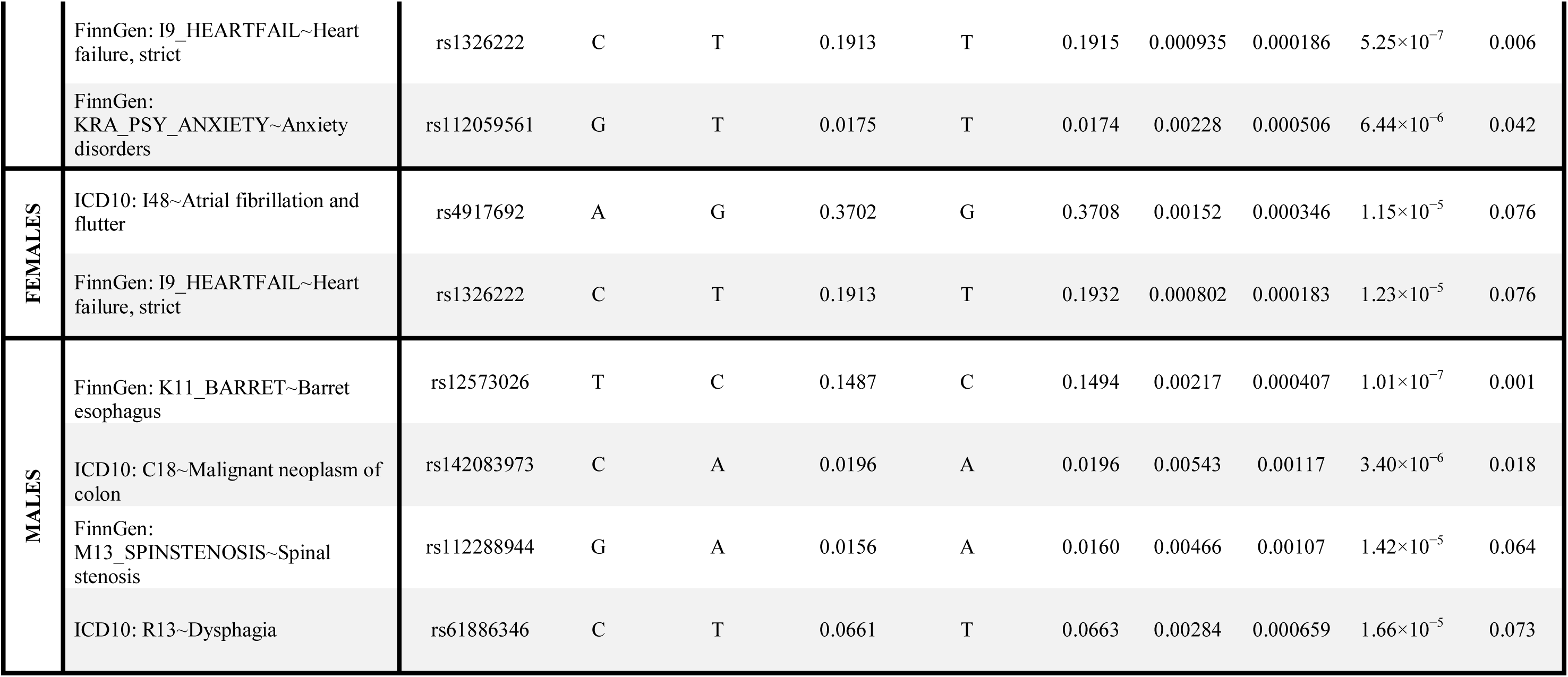
Allele frequencies of *TTR* and *RBP4* significant variants identified. Ref = reference allele on the forward strand; alt = alternate allele (not necessarily minor allele); AF = allele frequency; MAF = minor allele frequency in the n_complete_samples defined for associated phenotype.

## Notes

### Author Declarations

All relevant ethical guidelines have been followed and any necessary IRB and/or ethics committee approvals have been obtained.

Any clinical trials involved have been registered with an ICMJE-approved registry such as ClinicalTrials.gov and the trial ID is included in the manuscript.

